# Clinical and Genetic Associations of Deep Learning-Derived Cardiac Magnetic Resonance-Based Left Ventricular Mass

**DOI:** 10.1101/2022.01.09.22268962

**Authors:** Shaan Khurshid, Julieta Lazarte, James P. Pirruccello, Lu-Chen Weng, Seung Hoan Choi, Amelia W. Hall, Xin Wang, Samuel Friedman, Victor Nauffal, Kiran J. Biddinger, Krishna G. Aragam, Puneet Batra, Jennifer E. Ho, Anthony A. Philippakis, Patrick T. Ellinor, Steven A. Lubitz

**Affiliations:** Cardiovascular Research Center, Massachusetts General Hospital, Boston, Massachusetts, USA; Cardiovascular Disease Initiative, Broad Institute of Harvard and the Massachusetts Institute of Technology, Cambridge, Massachusetts, USA; Demoulas Center for Cardiac Arrhythmias, Massachusetts General Hospital, Boston, Massachusetts, USA; Division of Cardiology, Massachusetts General Hospital, Boston, Massachusetts, USA; Data Sciences Platform, Broad Institute of Harvard and the Massachusetts Institute of Technology, Cambridge, Massachusetts, USA; Division of Cardiology, Brigham and Women’s Hospital, Boston, Massachusetts, USA

**Keywords:** machine learning, left ventricular hypertrophy, genetics

## Abstract

Increased left ventricular (LV) mass (LVM) and LV hypertrophy (LVH) are risk markers for adverse cardiovascular events, and may indicate an underlying cardiomyopathy. Cardiac magnetic resonance (CMR) is the gold standard for LVM estimation, but is challenging to obtain at scale, which has limited the power of prior genetic analyses. In the current study, we performed a genome-wide association study (GWAS) of CMR-derived LVM indexed to body surface area (LVMI) estimated using a deep learning algorithm within nearly 50,000 participants from the UK Biobank. We identified 12 independent associations (1 known at *TTN* and 11 novel) meeting genome-wide significance, implicating several candidate genes previously associated with cardiac contractility and cardiomyopathy. Greater CMR-derived LVMI was associated with higher risk of incident dilated (hazard ratio [HR] 2.58 per 1-SD increase, 95% CI 2.10-3.17) and hypertrophic (HR 2.62, 95% CI 2.09-3.30) cardiomyopathies. A polygenic risk score (PRS) for LVMI was also associated with incident hypertrophic cardiomyopathy within a separate set of UK Biobank participants (HR 1.12, 95% CI 1.01-1.12) and among individuals in an external Mass General Brigham dataset (HR 1.18, 95% CI 1.01-1.37). In summary, using CMR-derived LVM available at scale, we have identified 12 common variants associated with LVMI (11 novel) and demonstrated that both CMR-derived and genetically determined LVMI are associated with risk of incident cardiomyopathy.

**Journal Subject Terms:** machine learning, left ventricular hypertrophy, genetics

## Introduction

Left ventricular hypertrophy (LVH) is defined as pathologically increased left ventricular mass (LVM)^1^ and is associated with increased risk of cardiovascular events including heart failure (HF),^1–3^ stroke,^1^ atrial fibrillation (AF),^4^ and sudden cardiac death.^5^ Increased LVM is also a hallmark of certain primary cardiomyopathies such as hypertrophic cardiomyopathy (HCM) and some dilated cardiomyopathies (DCM). Although LVM can be estimated using 12-lead electrocardiograms or echocardiography, cardiac magnetic resonance (CMR) offers more accurate and reproducible quantification of LVM, and has therefore emerged as the gold standard for diagnosing LVH.^6^

Imaging-based estimation of LVM typically requires LV segmentation, which is usually performed manually and requires substantial time and expertise. As a result, genetic analyses of imaging-based LVM have been limited by modest sample sizes. Genome-wide association studies (GWAS) of echocardiography-based LVM identified a single susceptibility locus downstream of *SPCS3*.^7–9^ More recently, a genome-wide association study within 19,000 individuals^10^ identified significant variants in the gene *TTN* associated with CMR-based LVM.

We developed a validated deep learning approach to automate estimation of LVM using CMR images (Machine Learning for Health – Segmentation [ML4H_seg_]), which may increase power to detect genetic associations underlying CMR-derived LVM.^11^ In the current study we utilized ML4H_seg_ to estimate LVM using CMRs from nearly 50,000 participants in the UK Biobank. Given that body size is a major determinant of LV size and mass,^12^ we analyzed LVMI (i.e., LVM indexed by body surface area) in our primary analyses, and assessed unindexed LVM in secondary analyses. Our GWAS of LVMI identified 12 independent variants meeting genome-wide significance, including 11 novel associations. Using expression quantitative trait loci (eQTLs), transcriptome-wide association testing (TWAS), and tissue-specific expression level data, we propose several candidate genes, many of which have been previously associated with cardiac contractility and cardiomyopathy. We additionally develop a polygenic risk score (PRS) for LVMI, and demonstrate that both phenotypic and genetic LVMI are associated with incident cardiovascular diseases including cardiomyopathy.

## Methods

### Data availability

UK Biobank data are publicly available by application (www.ukbiobank.ac.uk). Mass General Brigham (MGB) data contain protected health information and cannot be shared publicly. Data processing scripts used to perform the analyses described herein are available at https://github.com/shaankhurshid/lvmass_gwas.

### Study populations

The discovery sample comprised the UK Biobank, a population-based prospective cohort of 502,629 participants recruited between 2006-2010 in the United Kingdom to investigate the genetic and lifestyle determinants of disease. The design of the cohort has been described previously.^13,14^ Briefly, approximately 9.2 million individuals aged 40-69 years living within 25 miles of the 22 assessment centers in England, Wales, and Scotland were invited, and 5.4% participated in the baseline assessment. Extensive questionnaire data, physical measures, and biological data were collected at recruitment, with ongoing data collection in large subsets of the cohort, including repeated assessments and multimodal imaging. At the time of the current analysis, over 450,000 individuals have genome-wide genotyping data available. All participants are followed up for health outcomes through linkage to national health-related datasets.

We utilized the MGB Biobank to replicate a LVMI PRS that we derived in the UK Biobank. The MGB Biobank is a biorepository comprising patients from a multi-institutional healthcare network spanning seven hospitals in the New England region of the United States. MGB Biobank participants are followed for health outcomes through linkage to electronic health record (EHR) data.

UK Biobank and MGB Biobank participants provided written informed consent. The UK Biobank was approved by the UK Biobank Research Ethics Committee (reference number 11/NW/0382) and the MGB Biobank by the MGB Institutional Review Board. Use of UK Biobank (application #17488) and MGB Biobank data were approved by the local MGB Institutional Review Board.

### Cardiac magnetic resonance acquisition

For all analyses, we included individuals who underwent CMR during a UK Biobank imaging assessment and whose bulk CMR data were available for download as of 04-01/2020 (**Figure 1**). The full CMR protocol of the UK Biobank has been described in detail previously.^15^ Briefly, all CMR examinations were performed in the United Kingdom on a clinical wide-bore 1.5 Tesla scanner (MAGNETOM Aera, Syngo Platform VD13A, Siemens Healthineers, Erlangen, Germany). All acquisitions used balanced steady-state free precession with typical parameters.

**Figure 1.**
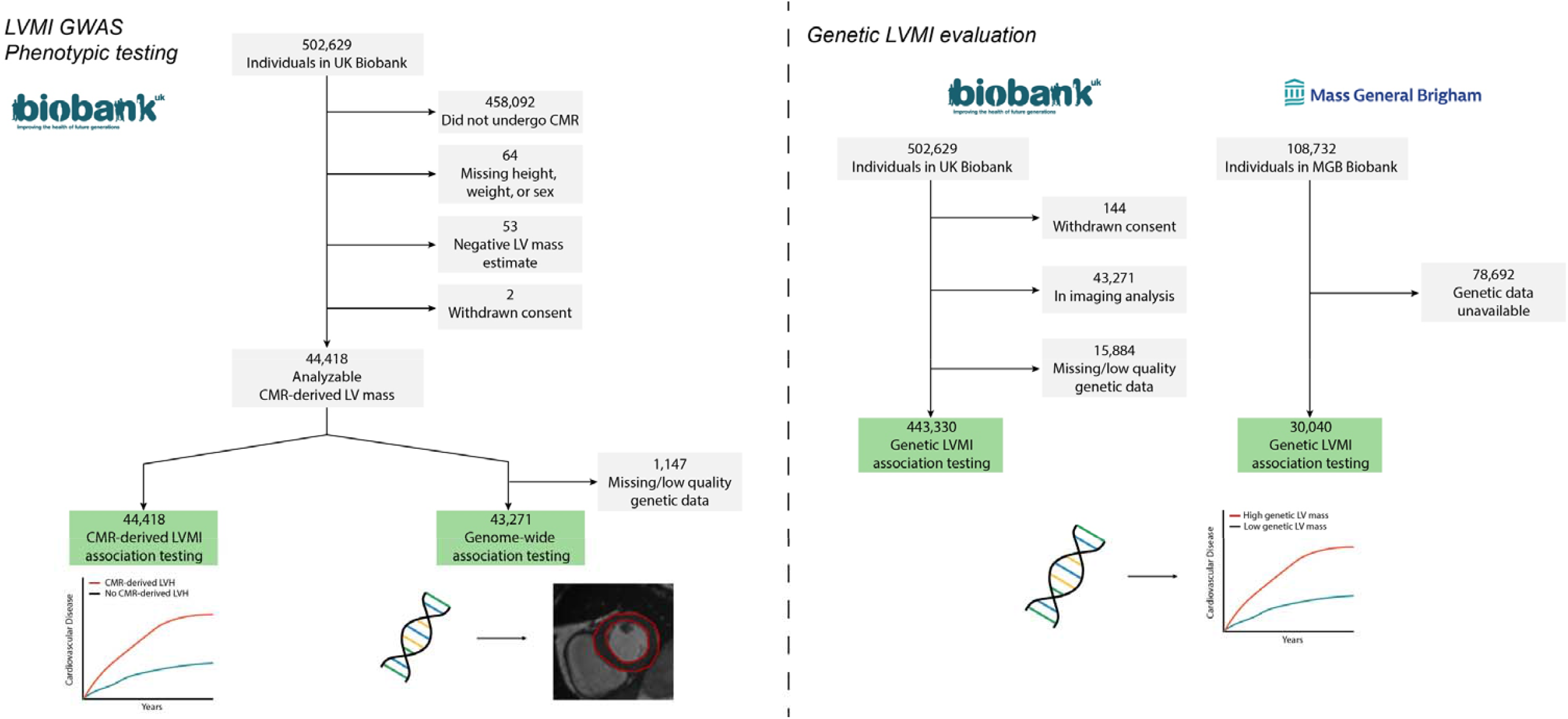
Overview of study design and flow We obtained CMR-derived LVM index in 44,418 individuals undergoing CMR imaging. We performed a genome-wide association study of CMR-derived LVMI and assessed for associations between CMR-derived LVMI and cardiovascular outcomes. Using GWAS results, we developed a polygenic risk score for LVMI, and applied it to 443,330 separate UK Biobank participants with genetic data to assess for associations between genetically determined LVMI and cardiovascular outcomes.

### Left ventricular mass estimation

We obtained CMR-derived LVM from all individuals with available CMR imaging using ML4H_seg_.^11^ Briefly, ML4H_seg_ identifies pixels corresponding to LV myocardium, which are then summed to estimate LV area and multiplied by slice thickness to estimate LV myocardial volume. LV myocardial volume is then multiplied by myocardial density (1.05 g/cm^3^) to yield LVM. As performed previously,^11^ LVM estimates were calibrated to the sex-specific sample means using manually labeled LVM measurements which were available within a subset of the UK Biobank sample (n=4,910). LVM estimates obtained using the described method have been shown to have very good correlation (Pearson *r* 0.86) and agreement (mean absolute error 10g) against manually labeled LVM in the UK Biobank.^11^ LVM estimates were indexed for body surface area using the DuBois formula to yield LVMI.^16^ A total of 53 (0.1%) individuals with estimated LVM values ≤0g were removed prior to analyses (**Figure 1**). The distribution of CMR-derived LVM is shown in **Supplemental Figure 1**.

### Genome-wide association study

To identify common genetic variation associated with CMR-derived LVM, we performed a GWAS of indexed LVM using BOLT-LMM.^17^ The model was adjusted for age at CMR acquisition, sex, array platform, and first five principal components of genetic ancestry. We excluded variants with minor allele frequency <1%. Associations were considered statistically significant at the standard genome-wide significance level (p=5×10^−8^). Lead single nucleotide polymorphisms (SNPs) were grouped into independent loci based on distance (±500kb), with conditional analyses performed to assess for independent signals within windows. Variants having suggestive (i.e., p<1×10^−6^) but not genome-wide significant associations were similarly tabulated. Genetic inflation was assessed by calculating the genomic control factor λ, inspecting quantile-quantile plots, and calculating the linkage disequilibrium score (LDSC) regression intercept.^18^ Observed scale heritability (*h*^*2*^) was estimated using the slope of LDSC regression. We assessed for independent signals within genome-wide significant loci by a) performing GWAS while conditioning on the imputed allele dosage of each lead SNP found in the primary GWAS (excluding insertion-deletion variants), and b) performing GWAS while conditioning on the top variant on chromosome 17 alone (rs6503451), to assess whether the additional variant located 914kb apart on chromosome 17 (rs199502, r^2^=0.37), was independent. The primary GWAS was performed among individuals of all genetic ancestries. In secondary analyses, we performed analogous GWAS restricted to individuals of European genetic ancestry, and also analyzed unindexed LV mass as the dependent variable. Further details of GWAS methods are provided in the **Supplemental Methods**.

### Bioinformatics and in silico functional analyses

We assessed whether genes within 500kb of lead SNPs were related to cardiac gene expression using GTEx^19^ version 8 *cis*-eQTL tissue data. To maximize power to detect potential candidate genes, as performed previously, we considered eQTLs for both atrial appendage (AA) and LV tissue data.^20,21^ We included lead variants as well as strong proxy variants (r^2^ ≥ 0.8). We also quantified tissue-specific expression levels from bulk RNA sequencing data from GTEx^19^ version 8. We evaluated the effects of predicted gene expression levels on LVMI by performing a transcriptome-wide association study (TWAS) using S-PrediXcan.^22^ GTEx genotypes and normalized expression data in AA and LV tissues provided in the software were used as training sets to develop the prediction models. Prediction models between each gene-tissue pair were developed using elastic net regression. In total, we tested 6,636 and 6,008 associations in AA and LV, respectively. The significance threshold for S-PrediXcan was therefore set at p=0.05/(6636+6008), or 3.95×10^−6^.

We prioritized likely candidate genes on the basis of proximity to the lead variant, eQTLs, TWAS, tissue-specific expression levels, and biologic plausibility based on previously reported data.

### Polygenic risk score development

To develop a PRS as a genetic instrument for CMR-derived LVMI, we applied a pruning and thresholding approach to our LVMI GWAS results. After removing insertion-deletion variants and strand ambiguous (i.e., A/T and C/G) variants to facilitate replication, we developed and tested four separate candidate PRS utilizing each combination of two thresholds used to define index SNPs (p=1×10^−6^ and p=1×10^−4^) and two thresholds used to prune proxy SNPs (r^2^=0.3 and r^2^=0.5). We then selected the PRS explaining the greatest variance in LVMI within the derivation set, which ultimately comprised a set of 475 variants (variance of LVMI explained=0.086; +3.60 g/m^2^ increase in LVMI per 1-standard deviation [1-SD] increase in PRS, p<0.01).

### Outcomes association testing

We assessed for associations between CMR-derived LVMI and incident AF, myocardial infarction, HF, ventricular arrhythmias, DCM, HCM, and implantable cardioverter-defibrillator (ICD) within participants with follow-up clinical data available after the imaging visit. We assessed for analogous associations using LVH, which was defined as LVMI >72g/m^2^ in men and >55 g/m^2^ in women,^23^ and alternatively as the sex-specific 90^th^ percentile of LVM.^1^ Diseases were defined using combinations of self-report and inpatient International Classification of Diseases, 9^th^ and 10^th^ revision codes (**Supplementary Table 1**). Start of follow-up was defined at the time of CMR acquisition and spanned until the earliest of an incident event, death, or last follow-up. The date of last follow-up was dependent upon the availability of linked hospital data, and was therefore defined as March 31, 2021 for participants enrolled in England (93.6%) and Scotland (6.1%), and February 28, 2018 for participants enrolled in Wales (0.3%).

We performed analogous association testing between the LVMI PRS and the same set of incident cardiovascular events among individuals in the UK Biobank that did not undergo CMR (n=443,330). Outcome and person-time definitions were similar, although start of follow-up was defined as the date of UK Biobank enrollment and blood sample collection. We also repeated association testing between the LVMI PRS and incident events in the independent MGB Biobank sample, using analogous models with person-time beginning at the date of blood sample collection and ending at an event, death, or last encounter in the EHR.

### Mendelian randomization analyses of blood pressure

As a form of validation of our LVM estimation, we sought to identify evidence of known causal associations between elevated blood pressure and increased LVM.^24^ We therefore conducted two-sample Mendelian randomization (MR) within individuals of genetic European ancestry in the UK Biobank sample. Genetic instruments were derived from a previous GWAS for systolic blood pressure (SBP) and diastolic blood pressure (DBP).^25^ Utilizing a 24 SNP instrument for SBP and a 26 SNP instrument for DBP, we prioritized inverse-variance weighted (IVW) meta-analyses of the effect of each SNP on CMR-derived LVMI (and LVM) divided by the effect of the same SNP on SBP or DBP. Weighted median and MR-Egger analyses were performed secondarily to address potential invalid instruments and directional pleiotropy. Further details of the MR analysis are provided in the **Supplemental Methods**.

### Statistical analysis

We tested associations between CMR-derived LVM and incident AF, myocardial infarction, HF, ventricular arrhythmias, DCM, HCM, and ICD using Cox proportional hazards regression with adjustment for sex and age at CMR acquisition. We fit analogous models using LVH (defined using the thresholds described above) and the LVMI PRS as the primary exposures. Models including the PRS were additionally adjusted for the first five principal components of genetic ancestry. Validity of the proportionality assumption was assessed using the Grambsch-Therneau test of correlation^26^ as well as visual inspection of smoothed fits to Schoenfeld residuals versus time. Where present, substantial deviations from proportional hazards (observed only for age, sex, and certain principal components of ancestry), were modeled by including interaction terms with strata of person-time.

Statistical analyses were performed using R v4.0 (packages ‘data.table’, ‘ggplot2’, ‘survival’, ‘prodlim’, ‘MendelianRandomization’).^27,28^ Except where otherwise noted, all two-tailed p-values <0.05 were considered statistically significant.

## Results

### Genome-wide association study of CMR-derived LVM

We conducted a multi-ancestry GWAS including 43,271 individuals (91% European ancestry) (**Table 1**). The analysis included a total of 9.9 million common variants imputed at an INFO score ≥0.30 and having minor allele frequency (MAF) ≥1%. The genomic control factor was 1.15 with a linkage disequilibrium score regression intercept of 1.00, consistent with polygenicity of the LVMI trait as opposed to inflation (**Supplementary Figure 2**). Observed scale *h*^*2*^ for LVMI was 0.26 (standard error [SE] 0.02).

**Table 1.**
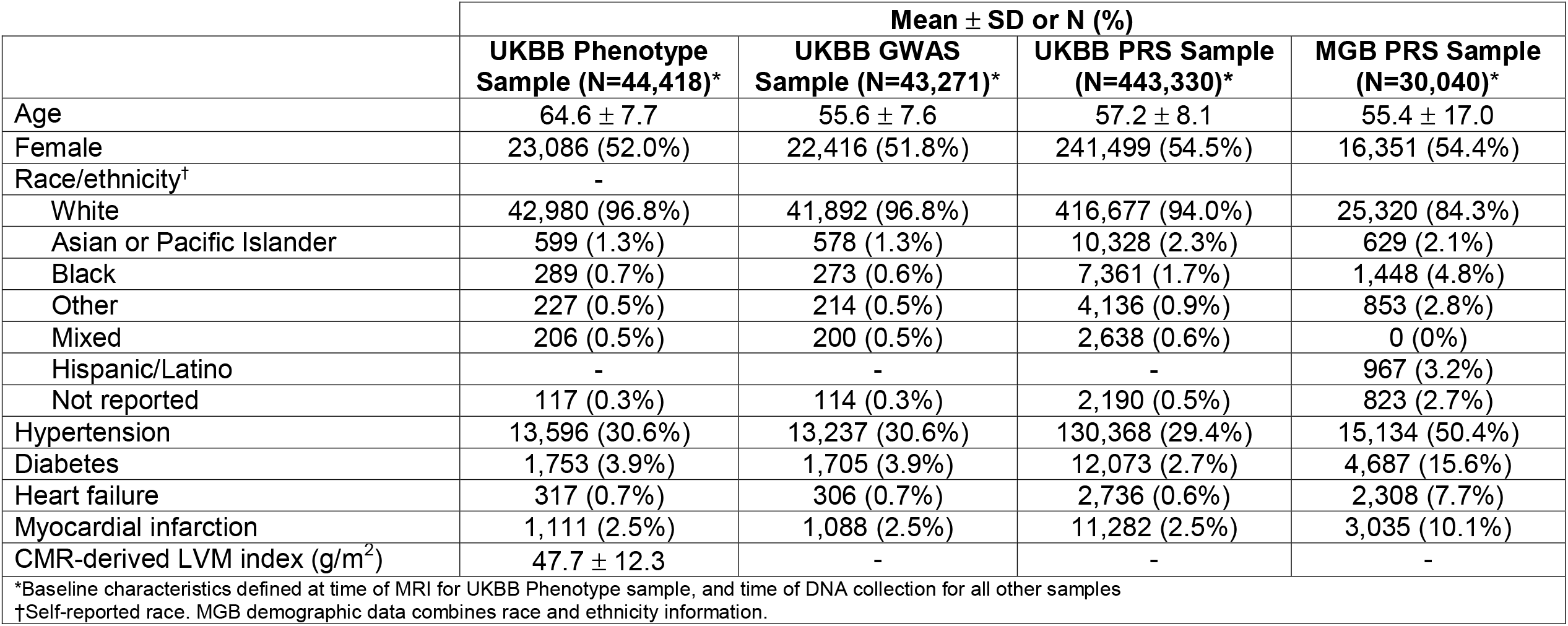
Baseline characteristics of study samples

| | Mean ± SD or N (%) | | | |
| --- | --- | --- | --- | --- |
|  | UKBB Phenotype Sample (N=44,418)* | UKBB GWAS Sample (N=43,271)* | UKBB PRS Sample (N=443,330)* | MGB PRS Sample (N=30,040)* |
| Age | 64.6 ± 7.7 | 55.6 ± 7.6 | 57.2 ± 8.1 | 55.4 ± 17.0 |
| Female | 23,086 (52.0%) | 22,416 (51.8%) | 241,499 (54.5%) | 16,351 (54.4%) |
| Race/ethnicity<sup>†</sup> | - |  |  |  |
| White | 42,980 (96.8%) | 41,892 (96.8%) | 416,677 (94.0%) | 25,320 (84.3%) |
| Asian or Pacific Islander | 599 (1.3%) | 578 (1.3%) | 10,328 (2.3%) | 629 (2.1%) |
| Black | 289 (0.7%) | 273 (0.6%) | 7,361 (1.7%) | 1,448 (4.8%) |
| Other | 227 (0.5%) | 214 (0.5%) | 4,136 (0.9%) | 853 (2.8%) |
| Mixed | 206 (0.5%) | 200 (0.5%) | 2,638 (0.6%) | 0 (0%) |
| Hispanic/Latino | - | - | - | 967 (3.2%) |
| Not reported | 117 (0.3%) | 114 (0.3%) | 2,190 (0.5%) | 823 (2.7%) |
| Hypertension | 13,596 (30.6%) | 13,237 (30.6%) | 130,368 (29.4%) | 15,134 (50.4%) |
| Diabetes | 1,753 (3.9%) | 1,705 (3.9%) | 12,073 (2.7%) | 4,687 (15.6%) |
| Heart failure | 317 (0.7%) | 306 (0.7%) | 2,736 (0.6%) | 2,308 (7.7%) |
| Myocardial infarction | 1,111 (2.5%) | 1,088 (2.5%) | 11,282 (2.5%) | 3,035 (10.1%) |
| CMR-derived LVM index (g/m<sup>2</sup>) | 47.7 ± 12.3 | - | - | - |
| *Baseline characteristics defined at time of MRI for UKBB Phenotype sample, and time of DNA collection for all other samples |  |  |  |  |
| <sup>†</sup>Self-reported race. MGB demographic data combines race and ethnicity information. |  |  |  |  |

The GWAS initially revealed 12 candidate SNPs associated with CMR-derived LVMI at genome-wide significance (**Table 2, Figure 2**). Conditional analyses identified an additional variant on chromosome 2, and that the two variants on chromosome 17 located 914kb apart (r^2^=0.37) were not independent, ultimately resulting in 12 lead SNPs for LVMI. The SNP most strongly associated with LVMI (rs2255167, p=1.7×10^−26^) was located at the *TTN* locus on chromosome 2 and has been previously associated with LVM. *TTN* is highly expressed in LV tissue (**Supplementary Table 2**).^10^ The remaining loci (n=11) were novel, with many located at or proximate to genes implicated in arrhythmias, cardiomyopathy and cardiomyocyte function, including *FLNC, MYOZ1, MAPT, WNT, CLCN6*, and *SYNPO2L*. Regional association plots for each genome-wide significant SNP are shown in **Supplementary Figure 3**. Results for 16 additional variants having suggestive but not genome-wide significant associations, including rs3729989 near the *MYBPC3* gene (p=5.9×10^−8^), are shown in **Supplementary Table 3**. A secondary GWAS of unindexed LVM revealed 12 genome-wide significant SNPs, of which 6 overlapped with the primary LVMI GWAS, and a 7^th^ was a strong proxy (r^2^=0.87). Loci unique to analyses of unindexed LVM appeared primarily enriched for genes associated with body size (e.g., *FTO, HMGA2, GDF5*), although *FTO* has also been implicated in HF^29^ and *CDKN1A* has been associated with DCM in a recent multi-trait analysis^30^ (**Supplementary Table 4** and **Supplementary Figure 4**).

**Table 2.** Variants associated with CMR-derived left ventricular mass index in the mixed-ancestry GWAS

| rsID | Chr | Position (hg38) | Closest gene(s) | Function | Risk/alt allele | RAF | Beta | SE | P-value |
| --- | --- | --- | --- | --- | --- | --- | --- | --- | --- |
| rs143800963 | 1 | 11835418 | *CLCN6* | Intronic | C/A | 0.95 | 0.94 | 0.16 | $9.0 \times 10^{-9}$ |
| rs2255167* | 2 | 178693555 | *TTN* | Intronic | T/A | 0.81 | 0.99 | 0.09 | $1.7 \times 10^{-26}$ |
| rs10497529<sup>†</sup> | 2 | 178975161 | *CCDC141* | Missense | G/A | 0.96 | 1.28 | 0.20 | $2.3 \times 10^{-10}$ |
| - | 5 | 133066736 | *HSPA4* | Indel | CTT/C | 0.72 | 0.50 | 0.08 | $1.2 \times 10^{-9}$ |
| rs9388498 | 6 | 126552277 | *CENPW* | - | G/T | 0.81 | -0.57 | 0.10 | $2.2 \times 10^{-9}$ |
| rs34163229 | 10 | 73647154 | *SYNPO2L* | Missense | G/T | 0.86 | -0.61 | 0.11 | $8.5 \times 10^{-9}$ |
| rs621679 | 11 | 1881538 | *LSP1* | Missense | G/A | 0.63 | -0.43 | 0.08 | $2.4 \times 10^{-8}$ |
| rs28552516 | 12 | 121592356 | *KDM2B* | Intronic | C/T | 0.85 | -0.58 | 0.10 | $1.8 \times 10^{-8}$ |
| rs6598541 | 15 | 98727906 | *IGF1R* | Intronic | A/G | 0.36 | -0.43 | 0.08 | $2.8 \times 10^{-8}$ |
| rs56252725 | 16 | 14995819 | *PDXDC1* | Intronic | G/A | 0.75 | 0.53 | 0.09 | $1.4 \times 10^{-8}$ |
| rs6503451 | 17 | 45870981 | *MAPT* | Intronic | T/C | 0.67 | -0.53 | 0.08 | $5.8 \times 10^{-11}$ |
| rs199502<sup>‡</sup> | 17 | 46784981 | *WNT3* | Intronic | G/A | 0.79 | -0.56 | 0.09 | $6.8 \times 10^{-10}$ |
| rs62621197 | 19 | 8605262 | *ADAMTS10* | Missense | C/T | 0.96 | 1.15 | 0.20 | $1.2 \times 10^{-8}$ |

\*Locus previously reported for LVM<sup>10</sup> <sup>†</sup>Variant identified in conditional analysis conditioned on lead SNPs (beta, standard error, and p-value are adjusted) <sup>‡</sup>Association no longer observed in analysis conditioned on rs6503451 Chr., chromosome; RAF, risk allele frequency; OR, odds ratio.

**Figure 2.**
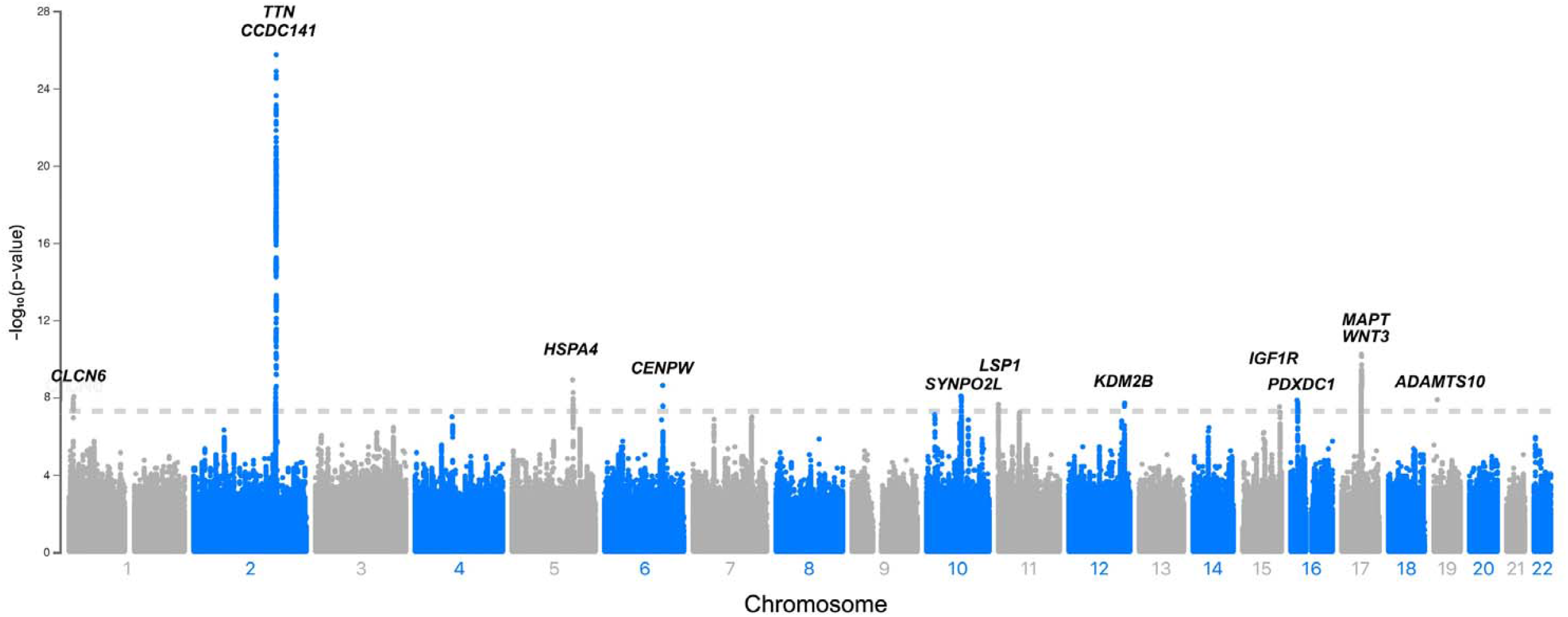
Manhattan plot of mixed-ancestry GWAS for CMR-derived LVM index Depicted across increasing chromosome (x-axis) are the results of the primary mixed-ancestry GWAS of left ventricular mass index. Variants meeting genome-wide significance (5×10^−8^, depicted by hashed horizontal line), are labeled by the closest gene to the lead variant.

In a GWAS restricted to individuals of European ancestry, 11 loci met genome-wide significance, of which 10 were shared with the primary GWAS (**Supplementary Table 5** and **Supplementary Figures 5-6**). The single locus unique to the European ancestry analysis was rs143973349, an insertion-deletion variant located near *FLNC*, a gene highly expressed in LV tissue and previously associated with familial hypertrophic, restrictive, and arrhythmogenic cardiomyopathies.^31–33^ This locus had a suggestive association with LVMI in the primary multi-ancestry GWAS (p=2.7×10^−7^).

### Bioinformatics and in silico functional analyses to determine candidate genes

In total, of the 12 independent lead SNPs, eight (or their proxies at r^2^ ≥ 0.8) were significant eQTLs in LV and/or AA tissue samples (**Figure 3**). The locus unique to the European ancestry subset also included eQTLs for LV and AA tissue. For a significant proportion of candidate genes, expression was identified in both LV and AA tissue samples. We then performed TWAS and identified 6 genes across 5 loci where predicted expression was associated with LVMI. Each of the genes implicated by TWAS was also an eQTL for either LV or AA (**Figure 3**). Detailed results of eQTL and TWAS analyses are shown in **Supplementary Table 2**.

**Figure 3.**
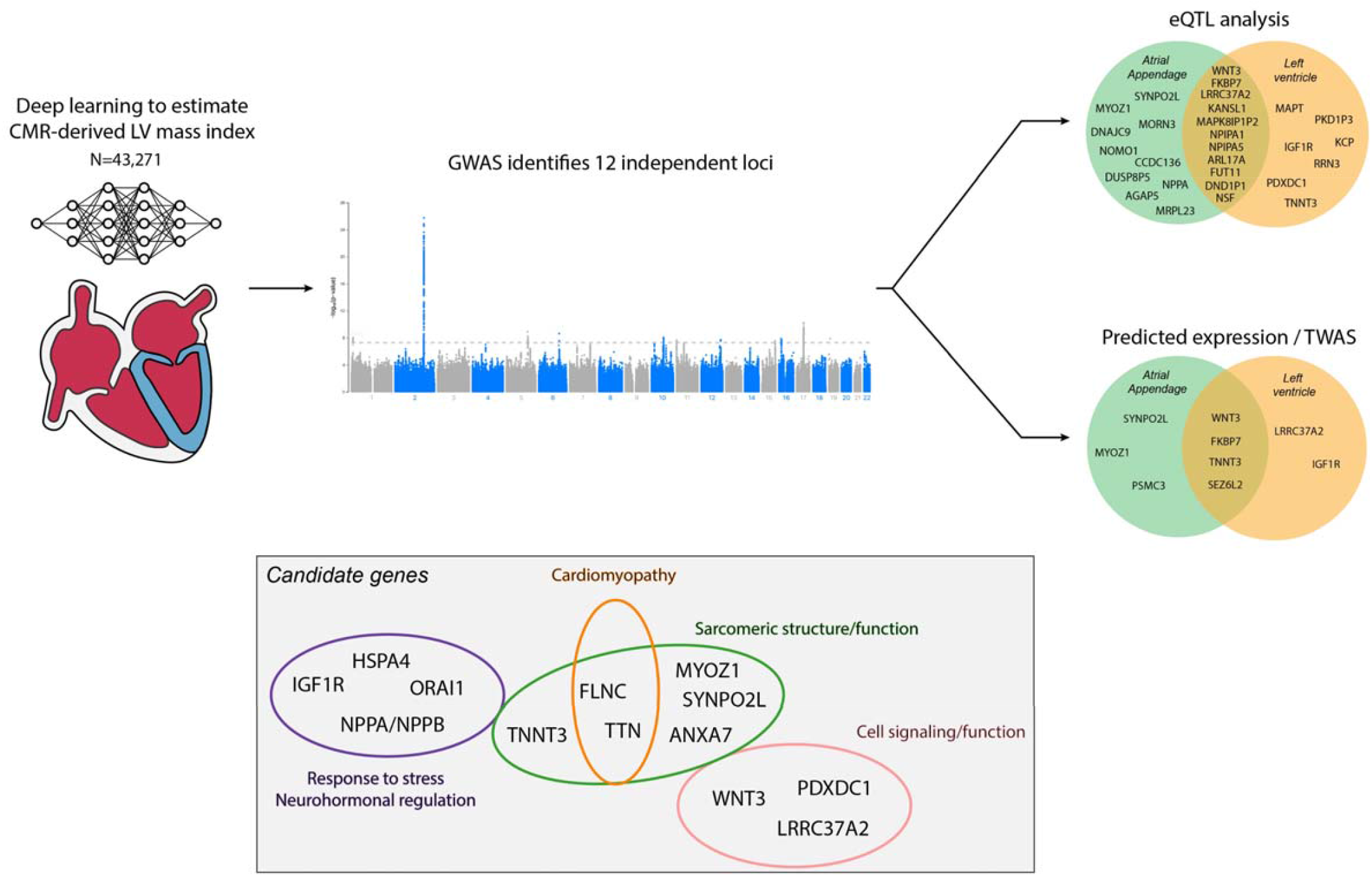
Candidate gene summary Depicted is a summary of study results. We used a deep learning algorithm to perform a GWAS of CMR-derived LVMI in 43,271 individuals, finding 12 independent loci associated with LVMI. Using proximity to lead variants, expression quantitative trait locus (eQTL) analysis, transcriptome-wide association studies (TWAS), and biologic plausibility, we identified candidate genes across the 12 loci. Candidate genes were enriched for genes involved in stress response and neurohormonal regulation, cardiac structure and cardiomyopathy, and cell signaling/function (gray box).

Probable candidate genes at each locus of interest are summarized in **Figure 3**. Specific genes implicated by multiple lines of evidence included *PDXDC1* near the rs56252725 locus, *IGF1R* near rs6598541, *SYNPO2L* and *MYOZ1* near rs34163229, *LRRC37A* near rs6503451, *WNT3* near rs199502, and *TNNT3* near rs149188822. For the locus detected only in the European ancestry subset, *KCP* near rs143973349 was also prioritized by multiple analyses. Selected genes prioritized solely based on strong biologic plausibility or previous associations with LVM included *TTN* near rs255167, *ORAI1* near rs28552516, and *FLNC* near rs143973349 (EUR only subset). Both *FLNC* and *TTN* have substantial expression in LV tissue (**Supplementary Table 2**).

### Associations between LVMI and cardiomyopathy

We assessed for associations between CMR-derived LVMI and incident cardiovascular disease. At a median follow-up of 2.7 years (Q1: 1.9, Q3: 4.1), greater LVMI was consistently associated with greater risk of multiple conditions, including AF, HF, DCM, and HCM (**Table 3**). CMR-derived LVH was strongly associated with incident DCM (HR 10.9, 95% CI 4.67-20.2), HCM (HR 9.26, 95% CI 3.20-26.8), and ICD insertion (HR 8.42, 95% CI 3.82-18.6). Cumulative risk of events stratified by presence versus absence of CMR-derived LVH is depicted in **Figure 4**.

**Table 3.** Associations between cardiac magnetic resonance-derived indexed LVM and incident cardiovascular disease

|  | N events / N total* | Follow-up,<br>years (Q1,Q3) | Hazard ratio for covariate (95% CI)<sup>†</sup> |  |  |
| --- | --- | --- | --- | --- | --- |
|  |  |  | LVMi (per 1 SD) | LVH | LVH (90<sup>th</sup> percentile) |
| Atrial fibrillation | 559 / 43045 | 2.7 (1.9, 4.1) | 1.46 (1.36-1.57) | 2.53 (2.02-3.17) | 2.53 (2.02-3.17) |
| Myocardial infarction | 299 / 43265 | 2.7 (1.9, 4.1) | 1.32 (1.19-1.47) | 1.57 (1.09-2.27) | 1.53 (1.10-2.13) |
| Heart failure | 267 / 44059 | 2.7 (1.9, 4.1) | 1.86 (1.70-2.04) | 4.96 (3.78-6.50) | 4.11 (3.15-5.35) |
| Ventricular arrhythmias | 110 / 44241 | 2.7 (1.9, 4.1) | 1.52 (1.30-1.78) | 2.83 (1.74-4.61) | 2.35 (1.47-3.75) |
| Dilated cardiomyopathy | 22 / 44346 | 2.7 (1.9, 4.1) | 2.58 (2.10-3.17) | 10.9 (4.67-20.2) | 11.4 (4.91-26.4s) |
| Hypertrophic cardiomyopathy | 14 / 44344 | 2.7 (1.9, 4.1) | 2.62 (2.09-3.30) | 9.26 (3.20-26.8) | 6.92 (2.40-20.0) |
| Implantable defibrillator | 26 / 44376 | 2.7 (1.9, 4.1) | 2.32 (1.89-2.84) | 8.42 (3.82-18.6) | 9.53 (4.41-20.6) |
| *Hazard ratios obtained using Cox proportional hazards models adjusted for age and sex<br>†N includes all individuals without the prevalent condition at the time of CMR acquisition<br>CI = confidence interval, LVMi = left ventricular mass index, LVH = left ventricular hypertrophy, Q1 = quartile 1, Q3 = quartile 3, SD = standard deviation |  |  |  |  |  |

**Figure 4.**
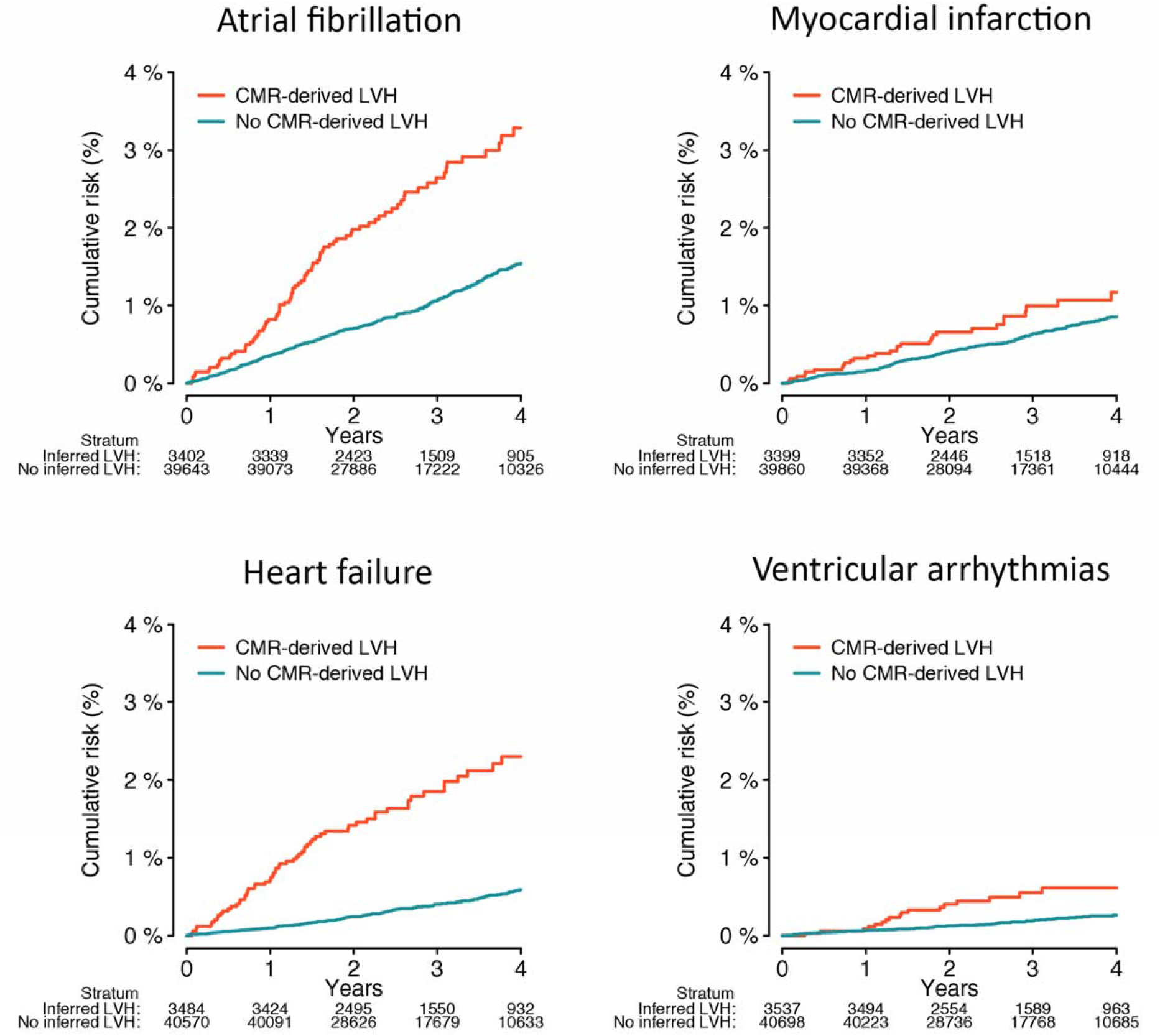
Kaplan-Meier plots of the association between CMR-derived LVH and incident cardiovascular disease Plots depicting the cumulative risk of atrial fibrillation (top left), heart failure (top right), myocardial infarction (bottom left), and ventricular arrhythmias (bottom right), stratified by the presence (orange) versus absence (teal) of CMR-derived LVH. LVH was defined as LVM index (LVMI) >72g/m^2^ in men and >55 g/m^2^ in women.^23^ The number at risk within each stratum over time is depicted below each plot.

We next evaluated associations between LVMI genetic risk and incident outcomes. In a set of UK Biobank participants separate from the GWAS sample (n= 443,330), a greater LVMI PRS was associated with higher risk of multiple incident conditions including AF, HF, DCM, HCM, and ICD (**Table 4**). In the independent MGB sample (n=30,040), the LVMI PRS was again associated with incident HCM and ICD (**Table 4**). In models of incident HCM risk, the relative hazard of HCM increased log-linearly with greater CMR-derived LVMI as well as LVMI PRS, with similar effect sizes in both the UK Biobank and MGB (**Figure 5**). Disease association results were similar in analyses restricted to individuals of European ancestry (**Supplementary Table 6**).

**Table 4.** Associations between LVMI PRS and incident disease

|  |  |  | Hazard ratio for covariate (95% CI)* |  |  |
| --- | --- | --- | --- | --- | --- |
|  | N events / N total† | Follow-up, yrs (Q1,Q3) | PRS (per 1 SD) | PRS (90<sup>th</sup> percentile) | PRS (95<sup>th</sup> percentile) |
| **UK Biobank** |  |  |  |  |  |
| Atrial fibrillation | 25050 / 435921 | 11.8 (11.0, 12.6) | 1.02 (1.00-1.03) | 1.05 (1.00-1.09) | 1.06 (1.00-1.12) |
| Myocardial infarction | 13408 / 432048 | 11.8 (11.0, 12.6) | 1.02 (1.00-1.04) | 1.08 (1.02-1.14) | 1.09 (1.02-1.18) |
| Heart failure | 13544 / 440594 | 11.9 (11.0, 12.6) | 1.04 (1.02-1.06) | 1.11 (1.02-1.13) | 1.07 (0.99-1.15) |
| Ventricular arrhythmias | 4881 / 442299 | 11.9 (11.1, 12.6) | 1.06 (1.03-1.09) | 1.15 (1.05-1.26) | 1.21 (1.07-1.36) |
| Dilated cardiomyopathy | 1023 / 443017 | 11.9 (11.1, 12.6) | 1.09 (1.03-1.16) | 1.24 (1.03-1.50) | 1.25 (0.96-1.61) |
| Hypertrophic cardiomyopathy | 419 / 443154 | 11.9 (11.1, 12.6) | 1.12 (1.02-1.24) | 1.07 (0.78-1.48) | 1.18 (0.78-1.80) |
| Implantable defibrillator | 1444 / 443219 | 11.9 (11.1, 12.6) | 1.06 (1.01-1.12) | 1.12 (0.95-1.32) | 1.26 (1.02-1.56) |
| **Mass General Brigham** |  |  |  |  |  |
| Atrial fibrillation | 1332 / 25305 | 2.9 (2.0, 4.1) | 1.01 (0.95-1.07) | 1.15 (0.97-1.36) | 1.35 (1.08-1.68) |
| Myocardial infarction | 695 / 28437 | 3.2 (2.1, 4.6) | 0.99 (0.92-1.07) | 1.09 (0.86-1.39) | 1.18 (0.85-1.63) |
| Heart failure | 1075 / 25054 | 2.9 (2.0, 4.1) | 0.97 (0.92-1.04) | 1.12 (0.93-1.36) | 1.17 (0.90-1.52) |
| Ventricular arrhythmias | 944 / 26979 | 3.0 (2.0, 4.2) | 0.97 (0.91-1.04) | 1.01 (0.82-1.25) | 1.15 (0.87-1.52) |
| Dilated cardiomyopathy | 450 / 28577 | 3.0 (2.1, 4.2) | 1.05 (0.95-1.15) | 1.10 (0.82-1.49) | 1.33 (0.90-1.95) |
| Hypertrophic cardiomyopathy | 183 / 28720 | 3.0 (2.1, 4.2) | 1.18 (1.01-1.37) | 1.11 (0.69-1.79) | 1.07 (0.54-2.09) |
| Implantable defibrillator | 137 / 28283 | 3.0 (2.1, 4.2) | 1.03 (0.86-1.22) | 1.02 (0.57-1.81) | 2.06 (1.13-3.74) |
| *Hazard ratios obtained using Cox proportional hazards models adjusted for age, sex, and principal components 1-5 |  |  |  |  |  |
| †N includes all individuals without the prevalent condition at baseline |  |  |  |  |  |
| CI = confidence interval, PRS = polygenic risk score, Q1 = quartile 1, Q3 = quartile 3, SD = standard deviation |  |  |  |  |  |

**Figure 5.**
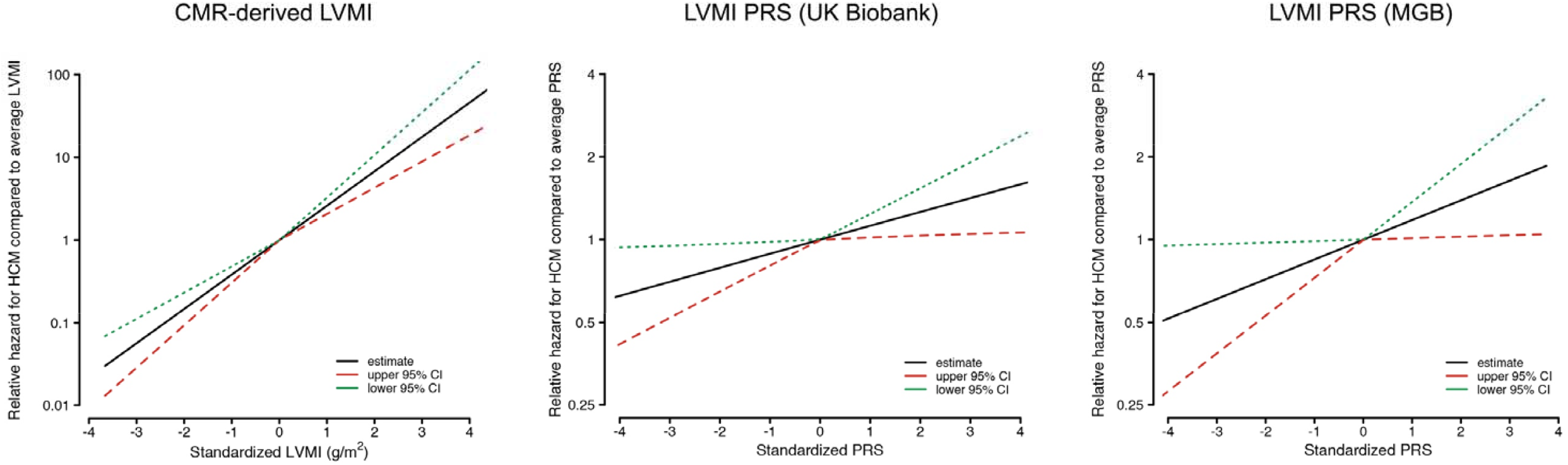
Association between CMR-derived and genetically predicted LVMI and incident HCM Depicted are plots showing the relative hazard of incident HCM as a function of increasing standardized CMR-derived LVM index (left), increasing standardized LVMI PRS in UK Biobank (middle) and increasing standardized LVMI PRS in Mass General Brigham (MGB, right). In each plot, the y-axis depicts the relative hazard of incident HCM compared to the hazard observed for individuals with an average LVMI (left) or average PRS value (middle and right), derived from Cox proportional hazards models adjusted for age and sex (left), and models adjusted for age, sex, and the first five principal components of genetic ancestry (middle and right). The relative hazard is plotted on the logarithmic scale. The functional form of the association was selected empirically using a penalized spline approach, in which the degrees of freedom for the penalized spline fit were chosen based on minimization of the corrected Akaike Information Criterion.^53^ In each case, an approximately linear relationship was chosen to model the relationship between the exposure and log hazard of HCM.

### Mendelian randomization

To assess for potential causal associations between blood pressure and CMR-derived LVMI, we performed MR analyses using genetic instruments for SBP and DBP. In an inverse variance weighted two-sample MR, a 1-SD increase in genetically mediated SBP was associated with a 0.203g increase in CMR-derived LVMI (95% CI, 0.076-0.33, p=0.002), and a 1-SD increase in genetically mediated DBP was associated with a 0.303g increase in CMR-derived LVMI (95% CI, 0.109-0.497, p=0.002). Weighted median two-sample MR analyses confirmed associations between both SBP and DBP and increased LVMI (p<5.34×10^−4^). MR-Egger analyses did not detect significant pleiotropy in any of the tested associations (intercept p>0.547). Associations were directionally similar but did not meet statistical significance for unindexed LVM (inverse variance weighted estimate: 0.265g per 1-SD increase in genetically mediated SBP, 95% CI -0.008-0.538, p=0.057 and 0.390g per 1-SD increase in genetically mediated DBP, 95% CI -0.041-0.821, p=0.08).

## Discussion

In the current study, we utilized a deep learning segmentation algorithm to perform GWAS of CMR-derived LVMI in nearly 50,000 individuals. Leveraging favorable statistical power and a rich imaging-based phenotype, we identified 12 independent loci associated with LVMI at genome-wide significance, including 11 associations that have not been previously reported. Downstream analyses prioritize several candidate genes, including multiple genes previously associated with cardiac structure and function, as well as cardiomyopathy. Importantly, both CMR-derived and genetically determined LVMI was associated with greater risk of incident cardiovascular events, and in particular incident HCM and DCM.

Our analyses suggest that common variants in cardiac structural and functional genes appear to be important determinants of LVM. Consistent with prior results,^10^ CMR-derived LVMI was strongly associated with variation at SNP rs2255167, located on the large sarcomeric protein *TTN. MYOZ1*, which was prioritized by both eQTL and TWAS and encodes a sarcomeric protein involved in the calcineurin signaling pathway, has been previously associated with both HF^29^ and AF.^34^ A mouse knockout of *MYOZ1* resulted in increased exercise capacity through activation of the nuclear factor of activated T-cells.^35^ Another gene prioritized by both eQTL and TWAS, *TNNT3*, encodes a troponin T isoform which is highly expressed in LV tissue. The *TNNT3* R63H variant has been shown to result in increased contractility in mouse skeletal muscle and is a known cause of the human disease Arthrogryposis (Type 2B2),^36^ which is characterized by limb contractures (i.e., excessive muscular contraction). *SYNPO2L*, an actin-related protein expressed in LV myocardium, has been previously associated with AF,^37^ HF,^38^ HCM,^30^ and voltage-duration product (i.e., a clinical indicator of LVH).^39^

Several of the candidate genes we identified prioritize neurohormonal regulation and response to physiologic stress as potential genetic determinants of LVMI. Specifically, lead variant rs143800963 is located on chromosome 1 within 20kb of *NPPA* and *NPPB*, genes that encode the natriuretic peptides Nppa and Nppb, respectively, with both proteins playing important roles in blood pressure regulation and salt homeostasis.^40^ Both Nppa and Nppb are constitutively expressed in ventricular myocardium and are also upregulated in response to stress.^41^ Knockout of *NPPB* in mice results in increased cardiac fibrosis in response to pressure overload.^42^ Conversely, cardiomyocyte-specific deletion of *ORAI1*, the product of which regulates calcium-induced calcium release, results in improved response to pressure overload, with preservation of normal calcium handling and LV systolic function as well as protection against angiotensin II-induced cardiac remodeling in adult myocardium.^43^ *IGFR1*, an eQTL for LV tissue and in which predicted expression in LV was associated with LVMI, encodes the insulin-like growth factor receptor 1, which has been implicated in organ growth and insulin resistance.^44^

Consistent with expectations, several LVMI candidate genes have previous links to cardiomyopathy and HF. The strongest association we observed with LVMI was at rs2255167, a variant located in *TTN*, in which mutations have been previously associated with familial cardiomyopathy^45^ and early-onset AF.^46^ One of the loci detected in the European ancestry sub-analysis (and suggestive in the primary analysis), *FLNC*, encodes filamin C, an actin-related protein associated with familial HCM,^32^ restrictive cardiomyopathy,^33^ and arrhythmogenic cardiomyopathy.^31^ A mouse knock-in of filamin C results in myofibrillar degeneration.^47^ *PPP3CB*, which encodes the signaling protein calcineurin, has been implicated in pathologic (as opposed to physiologic) cardiac hypertrophy.^48^ *FTO*, an obesity gene previously associated with HF,^29^ was associated with unindexed LV mass, but not for LVMI. Although not genome-wide significant (p=5.9×10^−8^), we also observed a suggestive association between LVMI and rs3729989 near the *MYBPC3* gene, which encodes a cardiac myosin-binding protein in which mutations are well-recognized causes of DCM and HCM.^49,50^

Importantly, we observed that both phenotypic and genetically determined LVMI were associated with substantially increased risks of incident cardiovascular events. Increased LVMI and LVH are consistently associated with an increased risk of HF.^2^ Indeed, we observed that greater LVMI was associated not only with greater risk of HF, but also incident DCM, HCM, and insertion of an ICD (a surrogate for cardiomyopathy or ventricular arrhythmias). Consistent with the notion that LVMI may be an endophenotype for certain cardiomyopathies, we observed that genetically determined LVMI – estimated using a 475-variant PRS – was consistently associated with greater risk of incident HCM within a separate set of UK Biobank participants as well as an external sample from the MGB healthcare system. Although recent work suggests that variants associated with risk of HCM and DCM tend to have opposite directions of effect,^30^ our findings support the notion that certain variants may increase risk of both conditions via pleiotropic effects on contractility and LV hypertrophy. Indeed, rs255167, the most significant association in our LVMI analysis, is within 100 base pairs of rs2042995, a variant associated with increased risk of both HCM and DCM in a prior multi-trait analysis.^30^ Overall, our findings provide evidence that the genetic variation underlying increased LVM may be clinically relevant, and may improve identification of individuals at greater risk of incident cardiomyopathy.

Our study has limitations. First, although our primary analysis was a mixed-ancestry GWAS, our sample is predominantly of European descent. As a result, our results may not generalize to individuals of other ancestries. Second, we used a deep learning model to estimate CMR-derived LVM, and therefore imperfect accuracy of estimations may have lead to reduced functional power to detect genetic associations. Nevertheless, we note that the model we utilized (ML4H_seg_) is very accurate, having a correlation of r=0.86 with hand-labeled CMR-derived LVM in the UK Biobank,^51^ and using MR analyses we were able to demonstrate evidence of known causal relationships between elevated blood pressure and increased indexed LVM.^24^ Third, our ability to assess for associations between CMR-derived LVMI and incident outcomes was limited by the amount of follow-up currently available after the imaging analysis. Fourth, generalizability of our findings may be affected by bias introduced by methods of enrollment, as the UK Biobank appears somewhat enriched for health and socioeconomic status as compared to the general population.^52^

In summary, we performed a GWAS of deep-learned CMR-derived LVM including nearly 50,000 individuals. We discovered 12 independent loci meeting genome-wide significance, including 11 that are novel. Using complementary downstream analyses, we identified multiple candidate genes, many of which are involved in cardiac structure and function, and several that have been previously implicated in cardiomyopathy. Both CMR-derived and genetically determined LVM were associated with incident HCM in independent datasets. Our findings add to our understanding of the common genetic variation underlying LVM and demonstrate the potential to use deep learning to define rich phenotypes at scale to empower clinically relevant biological discovery.

## Data Availability

UK Biobank data are publicly available by application (www.ukbiobank.ac.uk). MGB data contain protected health information and cannot be shared publicly. Data processing scripts used to perform the analyses described herein are available at https://github.com/shaankhurshid/lvmass_gwas.

