## Supplemental Material for "Clinical and Genetic Associations of Deep Learning-Derived Cardiac Magnetic Resonance-Based Left Ventricular Mass"

#### Supplementary Material

#### Index

**Methods.** Supplemental methods

*Genome-wide association study of LVMI*

We performed a genome-wide association study (GWAS) using CMR-derived LV mass index (LVMI), estimated using our deep learning segmentation model, as the variable of interest. The GWAS was performed using BOLT-LMM to perform a mixed-ancestry analysis by fitting a linear mixed model with segmentation model-based CMR-derived LVMI as the outcome of interest, with age at MRI, sex, array platform, and the first five principal components of genetic ancestry as covariates. The GWAS was performed among 32,328 individuals having undergone CMR imaging, after exclusion of individuals without genetic data meeting standard quality control metrics (e.g., no evidence of sex chromosome aneuploidy, outliers in heterozygosity and missing rates). Imputed variants were retained if the imputation information metric was ≥0.3. All variants with minor allele frequency <1% were excluded from the final analyses. Visual inspection of the resulting quantile-quantile plot did not suggest substantial inflation (**Supplementary Figure 2**).

*Mendelian randomization analyses of blood pressure*

Summary statistics from a prior GWAS provided effect estimates and standard errors for the association of each SNP with systolic and diastolic blood pressure.[1] Linear regression models were adjusted for age, sex, genotyping array, and the first ten principal components of genetic ancestry, to determine the beta coefficients and standard errors for the association of each SNP with the outcome (CMR-derived LVMI). These SNP-specific estimates were combined to conduct two-sample Mendelian randomization using the 'MendelianRandomization' package in R. Mendelian randomization analyses were limited to individuals of European genetic ancestry.

**Table 1**. Clinical factor definitions

| Phenotype | Data fields | Field names | Data codes | Data code definitions |
|---|---|---|---|---|
| **UK Biobank** | | | | |
| Atrial fibrillation | 20002 | Non-cancer illness code, self-reported | 1471, 1483 | Atrial fibrillation, Atrial flutter |
| Atrial fibrillation | 20004 | Operation code, self-reported | 1524 | Cardioversion |
| Atrial fibrillation | 41202<br>41204<br>40001<br>40002 | Diagnoses - main ICD10<br>Diagnoses – secondary ICD10<br>Underlying (primary) cause of death: ICD10,<br>Contributory (secondary) cause of death: ICD10 | I48, I48.0, I48.1, I48.2, I48.3, I48.4, I48.9 | Atrial fibrillation and flutter, Paroxysmal atrial fibrillation, Persistent atrial fibrillation, Chronic atrial fibrillation, Typical atrial flutter, Atypical atrial flutter, Atrial fibrillation and flutter, unspecified |
| Atrial fibrillation | 41203<br>41205 | Diagnosis - main ICD9<br>Diagnoses - secondary ICD9 | 4273 | Atrial fibrillation and flutter |
| Atrial fibrillation | 41200<br>41210 | Operative procedures – main OPCS<br>Operative procedures – secondary OPCS | K57.1, K62.1, K62.2, K62.3, K62.4, X50.1, X50.2 | Percutaneous transluminal ablation of atrioventricular node, Percutaneous transluminal ablation of pulmonary vein to left atrium conducting system, Percutaneous transluminal ablation of atrial wall for atrial flutter, Percutaneous transluminal ablation of conducting system of heart for atrial flutter NEC, Percutaneous transluminal internal cardioversion NEC, Direct current cardioversion, External cardioversion NEC |
| Diabetes | 2443 | Diabetes diagnosed by doctor | 1 | Yes |
| Diabetes | 20002 | Non-cancer illness code, self-reported | 1220, 1221, 1222, 1223 | Diabetes, Gestational diabetes, Type 1 diabetes, Type 2 diabetes |

| | | | | |
|---|---|---|---|---|
| Diabetes | 2986 | Insulin use within one year | 1 | Started insulin within one year diagnosis of diabetes - Yes |
| Diabetes | 6177 | Medication for cholesterol, blood pressure or diabetes | 3 | Insulin |
| Diabetes | 6153 | Medication for cholesterol, blood pressure, diabetes, or take exogenous hormones | 3 | Insulin |
| Diabetes | 41202 41204 40001 40002 | Diagnoses - main ICD10 Diagnoses – secondary ICD10 Underlying (primary) cause of death: ICD10, Contributory (secondary) cause of death: ICD10 | E10, E10.0, E10.1, E10.2, E10.3, E10.4, E10.5, E10.6, E10.7, E10.8, E10.9, E11, E11.0, E11.1, E11.2, E11.3, E11.4, E11.5, E11.6, E11.7, E11.8, E11.9, E12, E12.1, E12.8, E12.9, E13, E13.1, E13.2, E13.3, E13.5, E13.6, E13.7, E13.8, E13.9, E14, E14.0, E14.1, E14.2, E14.3, E14.4, E14.5, E14.6, E14.7, E14.8, E14.9 | Insulin-dependent diabetes mellitus, Insulin-dependent diabetes mellitus with coma, Insulin-dependent diabetes mellitus with ketoacidosis, Insulin-dependent diabetes mellitus with renal complications, Insulin-dependent diabetes mellitus with ophthalmic complications, Insulin-dependent diabetes mellitus with neurological complications, Insulin-dependent diabetes mellitus with peripheral circulatory complications, Insulin-dependent diabetes mellitus with other specified complications, Insulin-dependent diabetes mellitus with multiple complications, Insulin-dependent diabetes mellitus with unspecified complications, Insulin-dependent diabetes mellitus without complications, Non-insulin-dependent diabetes mellitus, Non-insulin-dependent diabetes mellitus - with coma, Non-insulin-dependent diabetes mellitus - with ketoacidosis, Non-insulin-dependent diabetes mellitus - with renal complications, Non-insulin-dependent diabetes mellitus - with ophthalmic complications, Non-insulin-dependent diabetes mellitus - with neurological complications, Non-insulin-dependent diabetes mellitus - with peripheral circulatory complications, Non-insulin-dependent diabetes mellitus - with other specified complications, Non-insulin-dependent diabetes mellitus - with multiple complications, Non-insulin-dependent diabetes mellitus - with unspecified complications, Non-insulin-dependent diabetes mellitus - without complications, Malnutrition-related diabetes mellitus, Malnutrition-related diabetes mellitus with ketoacidosis, Malnutrition-related diabetes mellitus with unspecified complications, Malnutrition-related diabetes mellitus without complications, Other specified diabetes mellitus, Other specified diabetes mellitus with ketoacidosis, Other specified diabetes mellitus with renal complications, Other specified diabetes mellitus with ophthalmic complications, Other specified diabetes mellitus with peripheral circulatory complications, Other specified diabetes mellitus with other specified complications, Other specified diabetes mellitus with multiple complications, Other specified diabetes mellitus with unspecified complications, Other specified diabetes mellitus without complications, Unspecified diabetes mellitus, Unspecified diabetes mellitus with coma, Unspecified diabetes mellitus with ketoacidosis, Unspecified diabetes mellitus with renal complications, Unspecified |

| | | | | |
|---|---|---|---|---|
| | | | | diabetes mellitus with ophthalmic complications, Unspecified diabetes mellitus with neurological complications, Unspecified diabetes mellitus with peripheral circulatory complications, Unspecified diabetes mellitus with other specified complications, Unspecified diabetes mellitus with multiple complications, Unspecified diabetes mellitus with unspecified complications, Unspecified diabetes mellitus without complications |
| Diabetes | 41203<br>41205 | Diagnosis - main ICD9 Diagnoses - secondary ICD9 | 2500, 25000, 25001, 25009, 2501, 25011, 25019, 2503, 2504, 2505, 25099 | Diabetes mellitus without mention of complication, Diabetes mellitus without mention of complication (adult-onset type), Diabetes mellitus without mention of complication (juvenile type), Diabetes mellitus without mention of compl. (adult/juvenile unspec.), Diabetes with ketoacidosis, Diabetes with ketoacidosis (juvenile type), Diabetes with ketoacidosis (adult/juvenile unspec.), Diabetes with renal manifestations, Diabetes with ophthalmic manifestations, Diabetes with neurological manifestations, Diabetes with unspecified complications (unspecified onset) |
| Heart Failure | 20002 | Non-cancer illness code, self-reported | 1076, 1079 | Heart failure/pulmonary oedema, Cardiomyopathy |
| Heart Failure | 41202<br>41204<br>40001<br>40002 | Diagnoses - main ICD10 Diagnoses – secondary ICD10 Underlying (primary) cause of death: ICD10, Contributory (secondary) cause of death: ICD10 | I11.0, I13.0, I13.2, I25.5, I42.0, I42.5, I42.8, I42.9, I50, I50.0, I50.1, I50.9 | Hypertensive heart disease with (congestive) heart failure, Hypertensive heart and renal disease with (congestive) heart failure, Hypertensive heart and renal disease with both (congestive) heart failure and renal failure, Hypertensive heart disease with (congestive) heart failure, Hypertensive heart and renal disease with (congestive) heart failure, Hypertensive heart and renal disease with both (congestive) heart failure and renal failure, Ischaemic cardiomyopathy, Dilated cardiomyopathy, Other restrictive cardiomyopathy, Other cardiomyopathies, Cardiomyopathy, unspecified, Heart failure, Congestive heart failure, Left ventricular failure, Heart failure, unspecified |
| Heart Failure | 41203<br>41205 | Diagnoses - main ICD9 Diagnoses – secondary ICD9 | 4254, 4280, 4281, 4289 | Other primary cardiomyopathies, Congestive heart failure, unspecified, Left heart failure, Heart failure, unspecified |
| Hypertension | 20002 | Non-cancer illness code, self-reported | 1065, 1072 | Hypertension, essential hypertension |
| Hypertension | 6150 | Vascular/heart problems diagnosed by doctor | 4 | High blood pressure |
| Hypertension | 41202<br>41204<br>40001<br>40002 | Diagnoses - main ICD10 | I10, I11, I11.0, I11.9, I12, I12.0, I12.9, I13, | Essential (primary) hypertension, Hypertensive heart disease, Hypertensive heart disease with (congestive) heart failure, Hypertensive heart disease without (congestive) heart failure, Hypertensive renal disease, Hypertensive renal disease with renal failure, Hypertensive renal disease without renal failure, Hypertensive heart and renal disease, |

| | | | | |
|---|---|---|---|---|
| | | Diagnoses – secondary ICD10 Underlying (primary) cause of death: ICD10, Contributory (secondary) cause of death: ICD10 | I13.0, I13.1, I13.2, I13.9, I15, I15.0, I15.1, I15.2, I15.8, I15.9 | Hypertensive heart and renal disease with (congestive) heart failure, Hypertensive heart and renal disease with renal failure, Hypertensive heart and renal disease with both (congestive) heart failure and renal failure, Hypertensive heart and renal disease, unspecified, Secondary hypertension, Renovascular hypertension, Hypertension secondary to other renal disorders, Hypertension secondary to endocrine disorders, Other secondary hypertension, Secondary hypertension, unspecified |
| Hypertension | 41203 41205 | Diagnoses - main ICD9 Diagnoses – secondary ICD9 | 401, 4010, 4011, 4019, 402, 4020, 4021, 4029, 403, 4030, 4031, 4039, 404, 4040, 4041, 4049, 405, 4050, 4051, 4059 | Essential hypertension, Essential hypertension, specified as malignant, Essential hypertension, specified as benign, Essential hypertension, not specified as malignant or benign, Hypertensive heart disease, Hypertensive heart disease, specified as malignant, Hypertensive heart disease, specified as benign, Hypertensive heart disease, not specified as malignant or benign, Hypertensive renal disease, Hypertensive renal disease, specified as malignant, Hypertensive renal disease, specified as benign, Hypertensive renal disease, not specified as malignant or benign, Hypertensive heart and renal disease, Hypertensive heart and renal disease, specified as malignant, Hypertensive heart and renal disease, specified as benign, Hypertensive heart and renal disease, not specified as malignant or benign, Secondary hypertension, Secondary hypertension, specified as malignant, Secondary hypertension, specified as benign, Secondary hypertension, not specified as malignant or benign |
| Myocardial infarction | 20002 | Non-cancer illness code, self-reported | 1075 | Heart attack/myocardial infarction |
| Myocardial infarction | 41202 41204 40001 40002 | Diagnoses - main ICD10 Diagnoses – secondary ICD10 Underlying (primary) cause of death: ICD10, Contributory (secondary) cause of death: ICD10 | I21, I21.0, I21.1, I21.2, I21.3, I21.4, I21.9, I22, I22.0, I22.1, I22.8, I22.9, I23, I23.0, I23.1, I23.2, I23.3, I23.4, I23.5, I23.6, I23.8, I24.1, I25.2 | Acute myocardial infarction, Acute transmural myocardial infarction of anterior wall, Acute transmural myocardial infarction of inferior wall, Acute transmural myocardial infarction of other sites, Acute transmural myocardial infarction of unspecified site, Acute subendocardial myocardial infarction, Acute myocardial infarction, unspecified, Subsequent myocardial infarction, Subsequent myocardial infarction of anterior wall, Subsequent myocardial infarction of inferior wall, Subsequent myocardial infarction of other sites, Subsequent myocardial infarction of unspecified site, Certain current complications following acute myocardial infarction, Hemopericardium as current complication following acute myocardial infarction, Atrial septal defect as current complication following acute myocardial infarction, Ventricular septal defect as current complication following acute myocardial infarction, Rupture of cardiac wall without hemopericardium as current complication following acute myocardial infarction, Rupture of chordae tendinae as current complication following acute myocardial infarction, Rupture of papillary muscle as current complication following acute myocardial infarction, Thrombosis of atrium, auricular appendage, and ventricle as current complication following acute myocardial infarction, Other current complications following acute myocardial infarction, Dressler's Syndrome, Old myocardial infarction |

| | | | | |
|---|---|---|---|---|
| Myocardial infarction | 41203<br>41205 | Diagnosis - main ICD9<br>Diagnoses - secondary ICD9 | 410, 410.0, 410.1, 410.2, 410.3, 410.4, 410.5, 410.6, 410.7, 410.8, 410.9, 411, 411.0, 411.1, 411.8, 412, 412.9, 429.79 | Myocardial infarction, Acute myocardial infarction of other anterior wall, episode of care unspecified, Acute myocardial infarction of inferolateral wall, episode of care unspecified, Acute myocardial infarction of inferoposterior wall, episode of care unspecified, Acute myocardial infarction of other inferior wall, episode of care unspecified, Acute myocardial infarction of other lateral wall, episode of care unspecified, True posterior wall infarction, episode of care unspecified, Subendocardial infarction, episode of care unspecified, Acute myocardial infarction of other specified sites, episode of care unspecified, Acute myocardial infarction of unspecified site, episode of care unspecified, Postmyocardial infarction syndrome, Intermediate coronary syndrome, Other acute and subacute complications of ischemic heart disease, Old myocardial infarction, Ill-defined descriptions and complications of heart disease, other |
| Ventricular arrhythmias | 41202<br>41204<br>40001<br>40002 | Diagnoses - main ICD10<br>Diagnoses – secondary ICD10<br>Underlying (primary) cause of death: ICD10,<br>Contributory (secondary) cause of death: ICD10 | I47.0, I47.2, I49.0, I49.3, I46.0, I46.1, I46.9 | Reentry ventricular arrhythmia, Ventricular tachycardia, Ventricular fibrillation and flutter, Ventricular premature depolarization, Cardiac arrest with successful resuscitation, Sudden cardiac death, so described, Cardiac arrest, unspecified |
| Ventricular arrhythmias | 41203<br>41205 | Diagnosis - main ICD9<br>Diagnoses - secondary ICD9 | 4271, 4274, 4275 | Paroxysmal ventricular tachycardia, Ventricular fibrillation and flutter, Cardiac arrest |
| Ventricular arrhythmias | 41200<br>41210 | Operative procedures – main OPCS<br>Operative procedures – secondary OPCS | K57.6, K64.1, X50.3, X50.4, X50.8, X50.9 | Percutaneous transluminal ablation of ventricular wall NEC, Percutaneous radiofrequency ablation of epicardium, Advanced cardiac pulmonary resuscitation, External ventricular defibrillation, Other specified external resuscitation, Unspecified external resuscitation |

**Table 2.** Bioinformatics and *in silico* functional analysis summary

| rsID | Chr | Position (hg38) | Closest gene | GTEx v8 eQTL LV* | GTEx v8 eQTL AA* | TWAS LV | TWAS AA | Plausible genes within 500kb | Prioritized candidate genes | Distance to lead SNP | LV expression level |
|---|---|---|---|---|---|---|---|---|---|---|---|
| ***Mixed-ancestry analysis*** | | | | | | | | | | | |
| rs143800963 | 1 | 11835418 | CLCN6 | - | NPPA | - | - | CLCN6, MTHFR, NPPA, NPPB | CLCN6 | 29322 | 7.28 |
| | | | | | | | | | NPPA | 10291 | 35.81 |
| | | | | | | | | | NPPB | 22046 | 26.77 |
| rs2255167 | 2 | 178693555 | TTN | FKBP7 | FKBP7 | FKBP7 | FKBP7 | TTN, FKBP7, CCDC141 | TTN† | 167566 | 66.76 |
| | | | | | | | | | FKBP7 | 229891 | 2.675 |
| rs10497529 | 2 | 178975161 | CCDC141 | - | - | - | - | TTN, CCDC141 | TTN† | 449172 | 66.76 |
| | | | | | | | | | CCDC141† | 145404 | 5.30 |
| - | 5 | 133066736 | HSPA4 | - | - | - | - | HSPA4, ZCCHC10 | HSPA4 | 14723 | 25.89 |
| rs9388498 | 6 | 126552277 | CENPW | - | - | - | - | CENPW | - | 212162 | 0.42 |
| rs34163229 | 10 | 73647154 | SYNPO2L | SYNPO2L | SYNPO2L | - | SYNPO2L | MYOZ1, PPP3CB, ANXA7, AGAP5, FUT11, SYNPO2L | SYNPO2L | 2273 | 84.32 |
| | | | | - | MYOZ1 | - | MYOZ1 | | MYOZ1 | 15542 | 1.06 |
| | | | | FUT11 | FUT11 | - | - | | ANXA7 | 272053 | 50.75 |
| | | | | - | AGAP5 | - | - | | PPP3CB | 210721 | 15.81 |
| | | | | - | DNAJC9 | - | - | | AGAP5 | 27133 | 1.74 |
| | | | | - | DUSP8P5 | - | - | | | | |
| | | | | - | AC073389.2 | - | - | | | | |
| | | | | - | - | - | - | | | | |
| rs621679 | 11 | 1881538 | LSP1 | TNNT3 | - | TNNT3 | - | LSP1, SYT8, TNNI2, PRR33, TNNT3, MRPL23, PSMC3 | TNNT3 | 38165 | 33.73 |
| | | | | - | MRPL23-AS1 | - | - | | | | |
| | | | | - | - | - | PSMC3 | | | | |
| rs28552516 | 12 | 121592356 | KDM2B | - | MORN3 | - | - | ORAI1, KDM2B, MORN3 | ORAI1† | 34194 | 4.91 |
| rs6598541 | 15 | 98727906 | IGF1R | IGF1R | - | IGF1R | - | IGF1R | IGF1R | 79367 | 5.71 |
| rs56252725 | 16 | 14995819 | PDXDC1 | PDXDC1 | - | PDXDC1 | - | PDXDC1, NOMO1 | PDXDC1 | 21228 | 15.96 |
| | | | | PKD1P3 | - | - | - | | NOMO1 | 162098 | 15.12 |
| | | | | NPIPA3 | - | - | - | | | | |

| | | | | | | | | | | | |
|---|---|---|---|---|---|---|---|---|---|---|---|
| | | | | NPIPA5 | NPIPA5 | - | - | | | | |
| | | | | RRN3 | - | - | - | | | | |
| | | | | - | NOMO1 | - | - | | | | |
| | | | | NPIPA1 | NPIPA1 | - | - | | | | |
| | | | | - | AC139256.1 | - | - | | | | |
| | | | | - | - | SEZ6L2 | - | | | | |
| rs6503451 | 17 | 45870981 | MAPT | MAPT | - | - | - | MAPT, LRRC37A, DND1P1, MAPK8IP1P2, KANSL1, ARL17A, WNT3, CRHR1 | KANSL1 | 158935 | 3.72 |
| | | | | LRRC37A4P | LRRC37A4P | - | - | | MAPT | 23546 | 4.75 |
| | | | | LRRC37A2 | LRRC37A2 | LRRC37A2 | - | | LRRC37A | 421752 | 0.12 |
| | | | | DND1P1 | DND1P1 | - | - | | CRHR1 | 86701 | 0.01 |
| | | | | AC126544.1 | AC126544.1 | - | - | | | | |
| | | | | MAPK8IP1P2 | MAPK8IP1P2 | - | - | | | | |
| | | | | LINC02210 | LINC02210 | - | - | | | | |
| | | | | KANSL1-AS1 | KANSL1-AS1 | - | - | | | | |
| | | | | AC005829.1 | AC005829.1 | - | - | | | | |
| | | | | AC005829.2 | AC005829.2 | - | - | | | | |
| | | | | ARL17A | ARL17A | - | - | | | | |
| | | | | - | NSF | - | - | | | | |
| | | | | WNT3 | WNT3 | - | - | | | | |
| | | | | - | RP11-707O23.5 | - | - | | | | |
| | | | | - | RP11-259G18.3 | - | - | | | | |
| | | | | - | RP11-259G18.1 | - | - | | | | |
| | | | | - | CRHR1-IT1 | - | - | | | | |
| rs199502 | 17 | 46784981 | WNT3 | WNT3 | WNT3 | WNT3 | WNT3 | NSF, WNT3, LRRC37A, KANSL1, ARL17A | WNT3 | 22475 | 0.41 |
| | | | | LRRC37A | LRRC37A | - | - | | KANSL1 | 755065 | 3.72 |
| | | | | ARL17A | ARL17A | - | - | | LRRC37A | 492248 | 0.12 |
| | | | | NSF | NSF | - | - | | | | |
| | | | | KANSL1 | - | - | - | | | | |
| rs62621197 | 19 | 8605262 | ADAMTS10 | - | - | - | - | ADAMTS10, MYO1F | ADAMTS10† | 25022 | 3.79 |

| | | | | | | | | | | | | |
|---|---|---|---|---|---|---|---|---|---|---|---|---|
| | | | | | | | | | | MYO1F† | 84484 | 1.85 |
| ***European-ancestry analysis*** | | | | | | | | | | | | |
| rs143973349 | 7 | 128866182 | ATP6V1F | KCP | - | KCP | - | | FLNC, ATP6V1F, KCP, CCDC136, OP1SW, IRF5 | FLNC† | 35776 | 291.60 |
| | | | | - | CCDC136 | - | - | | | KCP | 4140 | 0.17 |

*eQTLs determined using lead variants or proxy variants ($r^2 \geq 0.8$) using all-population or European-ancestry linkage disequilibrium maps from the 1,000 Genomes Project
†Gene prioritized at locus on the basis of strong biologic plausibility or previous association with LVM based on previous literature only

**Table 3.** Variants with suggestive associations with CMR-derived left ventricular mass index in the mixed-ancestry GWAS

| rsID | Chr | Position (hg 38) | Closest gene(s) | Function | Risk/alt allele | RAF | Beta | SE | P-value |
|---|---|---|---|---|---|---|---|---|---|
| rs140620427 | 2 | 66552028 | MEIS1 | Intronic | G/GAC | 0.64 | -0.40 | 0.08 | $4.8x10^{-07}$ |
| rs80197559 | 3 | 134708638 | EPHB1 | Intronic | A/T | 0.93 | 0.71 | 0.14 | $6.3x10^{-07}$ |
| - | 3 | 169759203 | ACTRT3 | - | CAA/C | 0.75 | 0.44 | 0.09 | $3.4x10^{-07}$ |
| rs36034102 | 4 | 80280894 | FGF5 | Intronic | G/T | 0.73 | -0.44 | 0.08 | $1.0x10^{-07}$ |
| rs7708324 | 5 | 148540531 | HTR4 | Intronic | A/G | 0.62 | -0.38 | 0.08 | $4.2x10^{-07}$ |
| rs234478 | 6 | 124040001 | NKAIN2 | Intronic | A/G | 0.52 | 0.39 | 0.07 | $1.4x10^{-07}$ |
| rs17172722 | 7 | 46580714 | IGFBP3 | - | C/T | 0.58 | -0.40 | 0.08 | $1.3x10^{-07}$ |
| rs13227216 | 7 | 128850499 | FLNC | Intronic | T/C | 0.81 | 0.50 | 0.09 | $1.0x10^{-07}$ |
| rs35886223 | 7 | 129370466 | AHCYL2 | Intronic | G/A | 0.67 | 0.41 | 0.08 | $3.1x10^{-07}$ |
| rs76540339 | 10 | 18538957 | CACNB2 | Intronic | A/C | 0.87 | 0.58 | 0.11 | $7.6x10^{-08}$ |
| rs12253621 | 10 | 70680191 | ADAMTS14 | Intronic | G/C | 0.74 | 0.44 | 0.09 | $2.1x10^{-07}$ |
| rs111555687 | 10 | 89819910 | KIF20B | - | T/C | 0.98 | 1.30 | 0.25 | $1.4x10^{-07}$ |
| rs3729989 | 11 | 47348490 | MYBPC3 | Missense | T/C | 0.87 | -0.59 | 0.11 | $5.9x10^{-08}$ |
| rs35443 | 12 | 115115073 | TBX3 | - | G/C | 0.61 | 0.40 | 0.08 | $1.6x10^{-07}$ |
| - | 14 | 53462409 | DDHD1 | - | AT/A | 0.76 | -0.44 | 0.09 | $3.5x10^{-07}$ |
| rs73468773 | 15 | 65274657 | PARP16 | Intronic | C/G | 0.97 | -1.02 | 0.20 | $6.2x10^{-07}$ |

Chr = chromosome; RAF = risk allele frequency

**Table 4.** Variants associated with unindexed left ventricular mass

| rsID | Chr | Position (hg 38) | Closest gene(s) | Function | Risk/alt allele | RAF | Beta | SE | P-value |
|---|---|---|---|---|---|---|---|---|---|
| rs2255167 | 2 | 178693555 | TTN | Intronic | T/A | 0.81 | 1.85 | 0.21 | $1.8 \times 10^{-19}$ |
| - | 5 | 133066736 | HSPA4† | Indel | CTT/C | 0.72 | 1.02 | 0.18 | $2.8 \times 10^{-08}$ |
| rs3176323 | 6 | 36679072 | CDKN1A† | Intronic | T/C | 0.69 | -0.97 | 0.18 | $4.4 \times 10^{-08}$ |
| rs9388498 | 6 | 126552277 | CENPW | - | G/T | 0.81 | -1.46 | 0.22 | $9.1 \times 10^{-12}$ |
| rs10878349 | 12 | 65933852 | HMGA2† | Intronic | A/G | 0.49 | 0.98 | 0.16 | $1.9 \times 10^{-09}$ |
| - | 13 | 49990968 | TRIM13† | Indel | ACT/A | 0.98 | -3.31 | 0.59 | $2.3 \times 10^{-08}$ |
| rs4985155 | 16 | 15035602 | PDXDC1 | Intronic | A/G | 0.66 | 0.95 | 0.17 | $3.2 \times 10^{-08}$ |
| - | 16 | 53778858 | FTO† | Indel | ATTTT/A | 0.60 | -1.03 | 0.17 | $8.6 \times 10^{-10}$ |
| rs6503451 | 17 | 45870981 | MAPT | Intronic | T/C | 0.67 | -1.04 | 0.18 | $7.2 \times 10^{-09}$ |
| rs199508‡ | 17 | 46781472 | WNT3 | Intronic | C/G | 0.24 | 1.06 | 0.19 | $3.7 \times 10^{-08}$ |
| rs62621197 | 19 | 8605262 | ADAMTS10† | Missense | C/T | 0.96 | 2.91 | 0.45 | $1.3 \times 10^{-10}$ |
| rs143384 | 20 | 35437976 | GDF5† | 5'-UTR | A/G | 0.59 | -0.95 | 0.17 | $2.1 \times 10^{-08}$ |

*Locus previously reported for LV mass[2]
†Variant association unique to unindexed LV mass
‡Association not observed in conditional analyses adjusting for rs6503451
Chr = chromosome; RAF = risk allele frequency

**Table 5.** Variants associated with CMR-derived left ventricular mass index in the European-ancestry GWAS

| rsID | Chr | Position (hg 38) | Closest gene(s) | Function | Risk/alt allele | RAF | Beta | SE | P-value |
|---|---|---|---|---|---|---|---|---|---|
| rs143800963 | 1 | 11835418 | CLCN6 | Intronic | C/A | 0.95 | 0.97 | 0.17 | $1.5 \times 10^{-08}$ |
| rs2255167* | 2 | 178693555 | TTN | Intronic | T/A | 0.81 | 0.99 | 0.10 | $2.9 \times 10^{-24}$ |
| - | 5 | 133066736 | HSPA4 | Indel | CTT/C | 0.72 | 0.48 | 0.09 | $3.6 \times 10^{-08}$ |
| rs9388498 | 6 | 126552277 | CENPW | - | G/T | 0.81 | -0.59 | 0.10 | $3.9 \times 10^{-09}$ |
| rs143973349† | 7 | 128866181 | ATP6V1F | Indel | TGG/T | 0.82 | 0.58 | 0.10 | $1.2 \times 10^{-08}$ |
| rs2177843 | 10 | 73650119 | SYNPO2L | Missense | C/T | 0.85 | -0.59 | 0.11 | $4.9 \times 10^{-08}$ |
| rs149188822 | 11 | 1873863 | LSP1 | Intronic | T/C | 0.58 | 0.45 | 0.08 | $2.8 \times 10^{-08}$ |
| rs28552516 | 12 | 121592356 | KDM2B | Intronic | C/T | 0.85 | -0.65 | 0.11 | $2.5 \times 10^{-09}$ |
| rs6503451 | 17 | 45870981 | MAPT | Intronic | T/C | 0.67 | -0.55 | 0.08 | $3.8 \times 10^{-11}$ |
| rs146122400 | 17 | 46704858 | NSF | Intronic | A/AGT | 0.80 | -0.59 | 0.10 | $2.3 \times 10^{-09}$ |
| rs62621197 | 19 | 8605262 | ADAMTS10 | Missense | C/T | 0.96 | 1.28 | 0.21 | $1.5 \times 10^{-09}$ |

*Locus previously reported
†Variant association unique to European ancestry analysis
Chr = chromosome; RAF = risk allele frequency

**Table 6.** Associations between LVMI PRS and incident disease in European ancestry subset

| | | | Hazard ratio for covariate (95% CI)* | | |
|---|---|---|---|---|---|
| **Disease** | **N events / N total†** | **Follow-up, yrs (Q1,Q3)** | **PRS (per 1 SD)** | **PRS (90th percentile)** | **PRS (95th percentile)** |
| ***UK Biobank*** | | | | | |
| Atrial fibrillation | 20289 / 380809 | 10.8 (10.0, 11.5) | 1.01 (1.00-1.02) | 1.02 (0.97-1.07) | 1.05 (0.99-1.12) |
| Myocardial infarction | 10427 / 377602 | 10.9 (10.1, 11.6) | 1.03 (1.01-1.05) | 1.07 (1.00-1.14) | 1.06 (0.97-1.15) |
| Heart failure | 10417 / 385146 | 10.9 (10.1, 11.6) | 1.04 (1.02-1.06) | 1.07 (1.00-1.14) | 1.11 (1.02-1.21) |
| Ventricular arrhythmias | 3814 / 386659 | 10.9 (10.1, 11.6) | 1.07 (1.03-1.10) | 1.20 (1.09-1.33) | 1.27 (1.12-1.45) |
| Dilated CM | 830 / 387310 | 10.9 (10.1, 11.6) | 1.11 (1.04-1.19) | 1.12 (0.90-1.39) | 1.29 (0.98-1.71) |
| Hypertrophic CM | 285 / 387449 | 10.9 (10.1, 11.6) | 1.21 (1.08-1.36) | 1.39 (0.99-1.95) | 1.67 (1.09-2.56) |
| ICD | 1175 / 388064 | 10.9 (10.1, 11.6) | 1.09 (1.03-1.16) | 1.30 (1.09-1.54) | 1.43 (1.14-1.79) |
| ***Mass General Brigham*** | | | | | |
| Atrial fibrillation | 1189 / 23606 | 3.0 (2.0, 4.1) | 1.03 (0.97-1.09) | 1.07 (0.89-1.29) | 1.09 (0.84-1.41) |
| Myocardial infarction | 593 / 23916 | 3.0 (2.0, 4.1) | 1.01 (0.94-1.10) | 1.15 (0.89-1.49) | 1.29 (0.92-1.80) |
| Heart failure | 941 / 21092 | 2.9 (2.0, 4.1) | 1.00 (0.93-1.06) | 1.11 (0.91-1.37) | 1.14 (0.86-1.52) |
| Ventricular arrhythmias | 839 / 22674 | 3.0 (2.0, 4.2) | 0.97 (0.91-1.04) | 1.10 (0.89-1.37) | 1.02 (0.75-1.40) |
| Dilated CM | 366 / 24111 | 3.0 (2.1, 4.3) | 1.04 (0.94-1.16) | 0.92 (0.65-1.30) | 0.64 (0.36-1.14) |
| Hypertrophic CM | 147 / 24230 | 3.0 (2.1, 4.3) | 1.14 (0.97-1.34) | 1.08 (0.64-1.81) | 0.97 (0.45-2.06) |
| ICD | 125 / 23990 | 3.0 (2.1, 4.3) | 0.99 (0.83-1.18) | 1.14 (0.65-1.99) | 1.17 (0.55-2.51) |

*Hazard ratios obtained using Cox proportional hazards models adjusted for age, sex, and PCs 1-5
†N includes all individuals without the prevalent condition at baseline

**Figure 1.** Distribution of CMR-derived LV mass

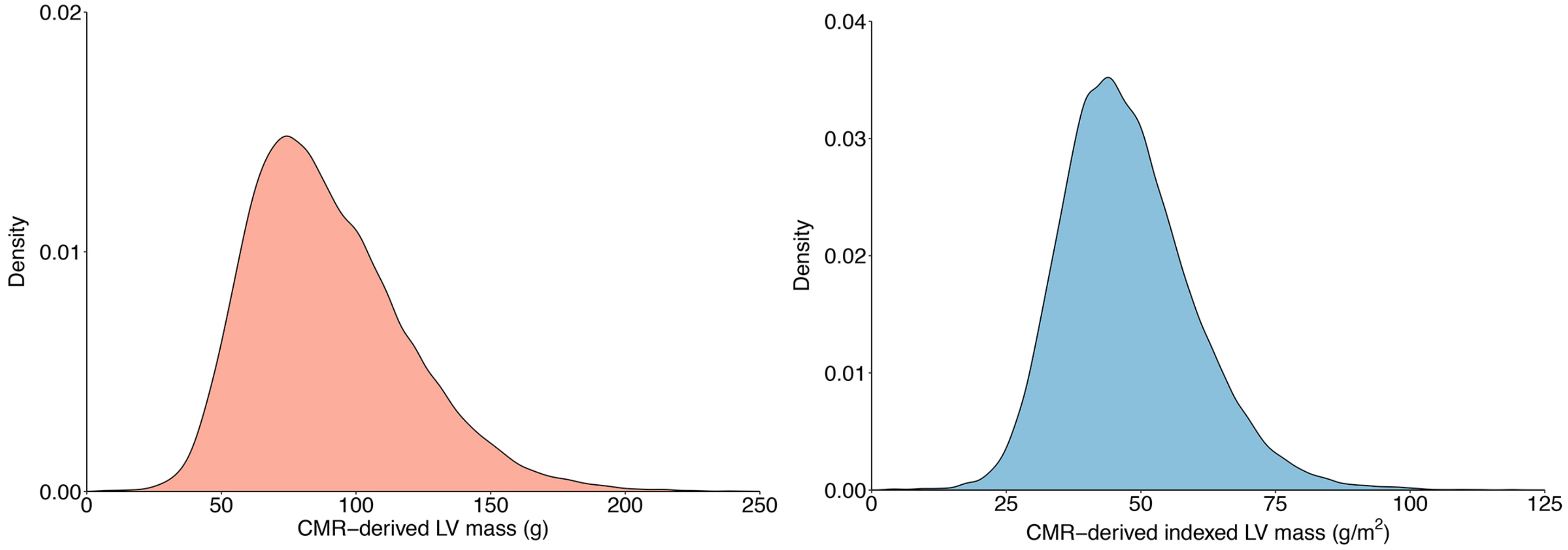

Depicted is the distribution of CMR-derived LV mass (LVM, left) and LV mass index (LVMI, right) within the UK Biobank phenotypic sample (N=44,418). The y-axis depicts the relative probability of an encountering a given value on the x-axis. Seven high outlying observations for LVM and six high outlying observations for LVMI are not depicted for graphical purposes.

**Figure 2.** Quantile-quantile plot of LVMI and LVM GWAS

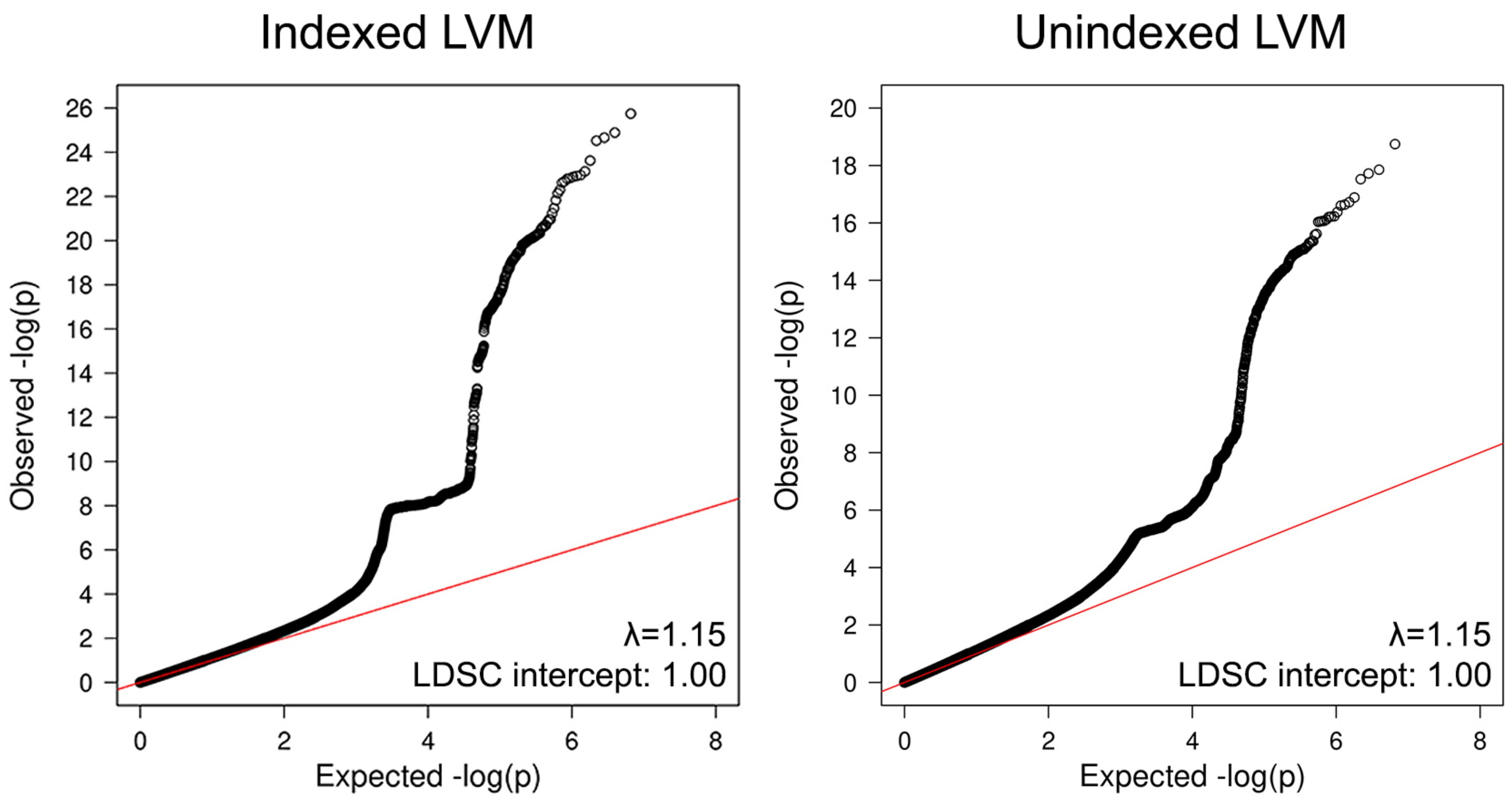

Depicted are quantile-quantile plots for the primary mixed-ancestry LVMI GWAS (left) and the unindexed LVM GWAS (right). The genomic control factor λ is depicted on the bottom right of each plot, as is the linkage disequilibrium score regression intercept, where a value close of one suggests the absence of inflation (and where an elevated λ is instead attributable to polygenicity).[3]

**Figure 3.** Regional association plots for genome-wide significant loci

rs143800963

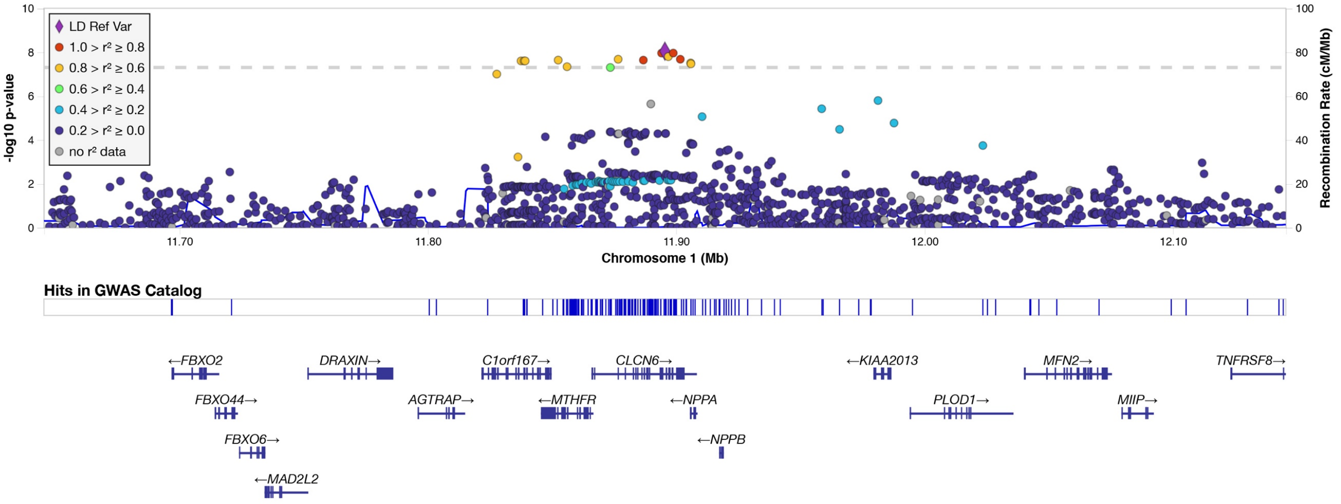

rs2255167

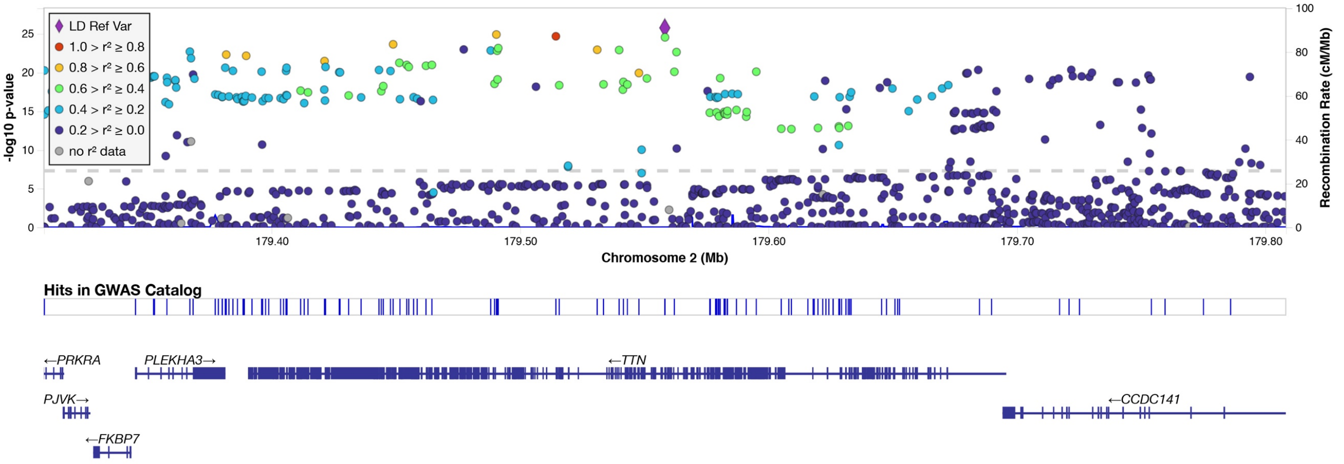

rs10497529

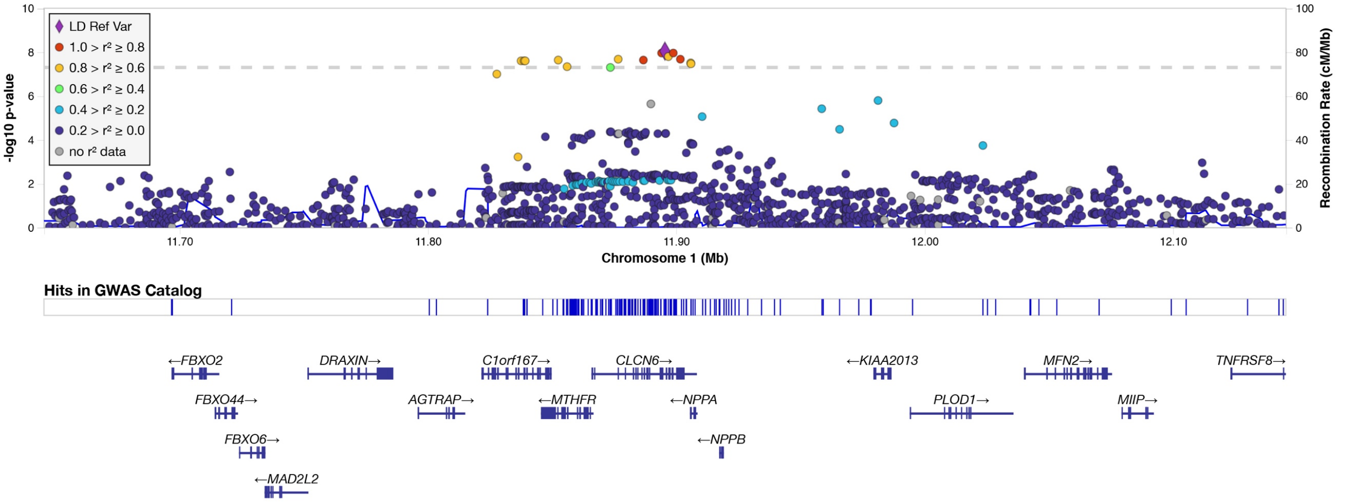

### 132402428:CTT_C

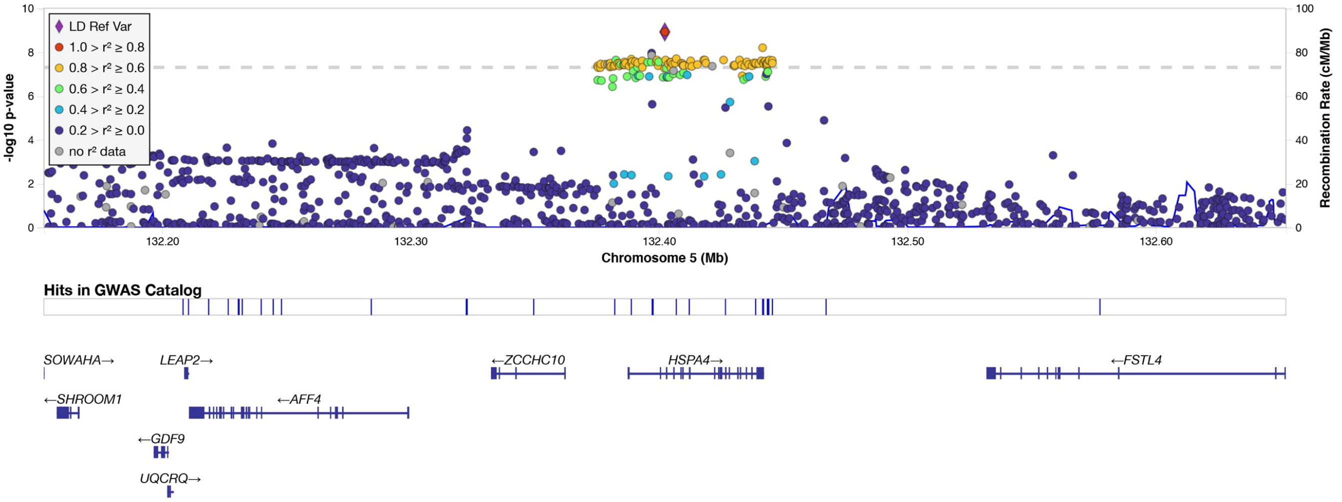

## rs9388498

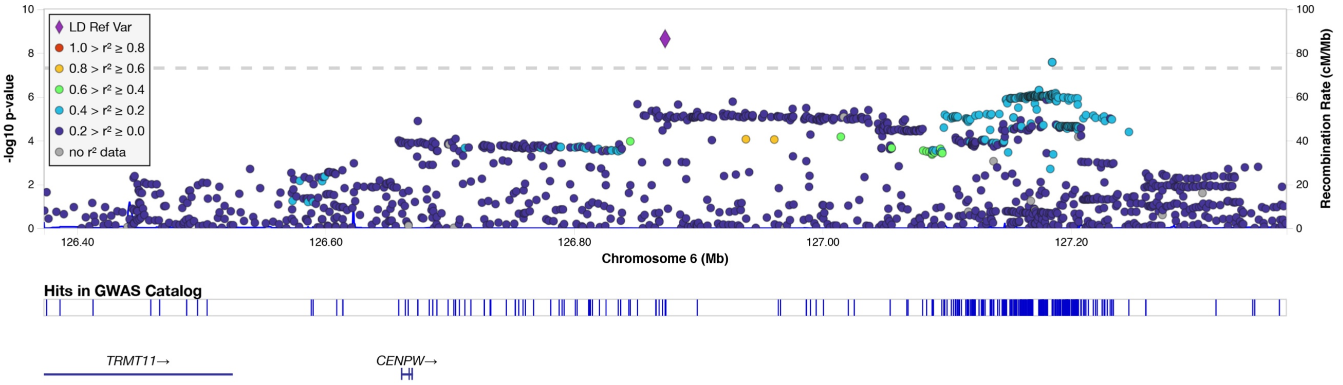

## rs34163229

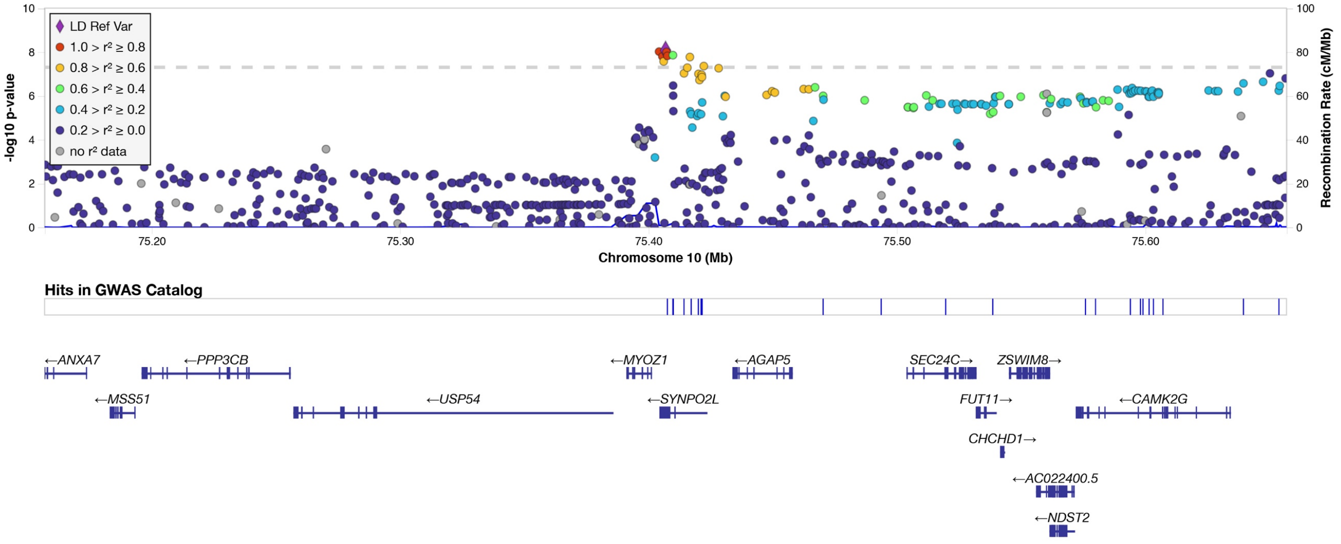

rs621679

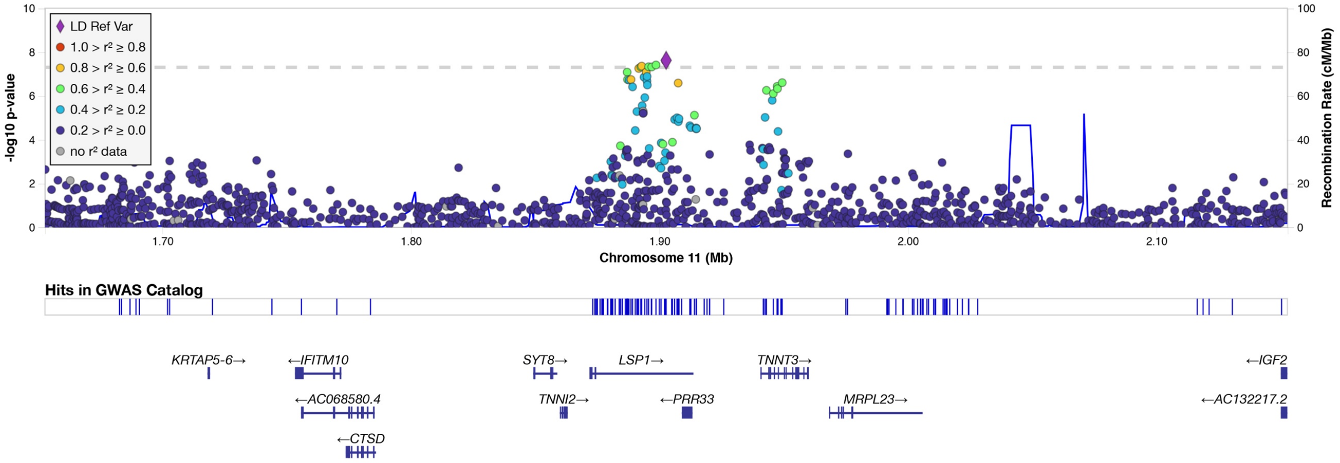

rs28552516

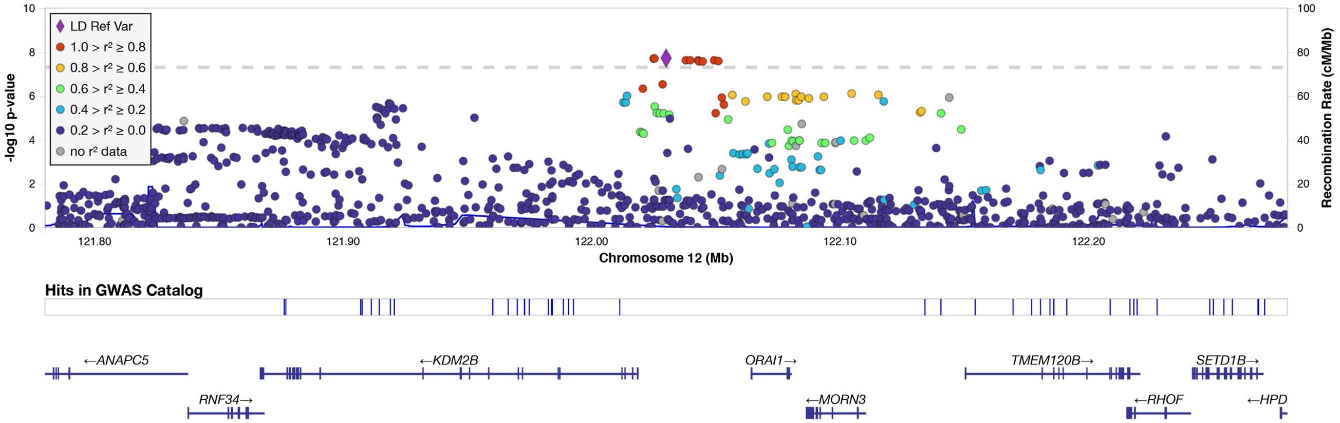

rs6598541

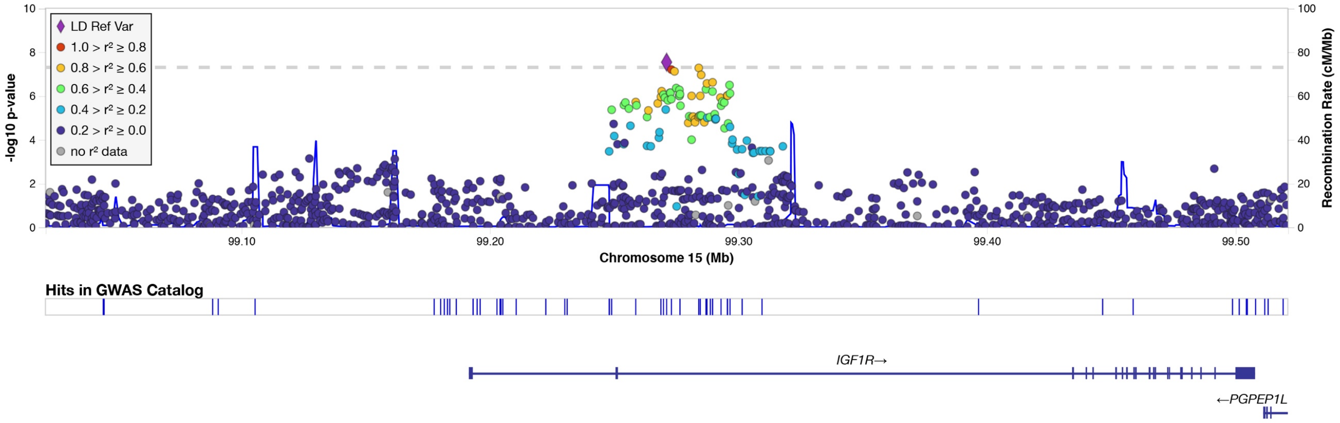

## rs56252725

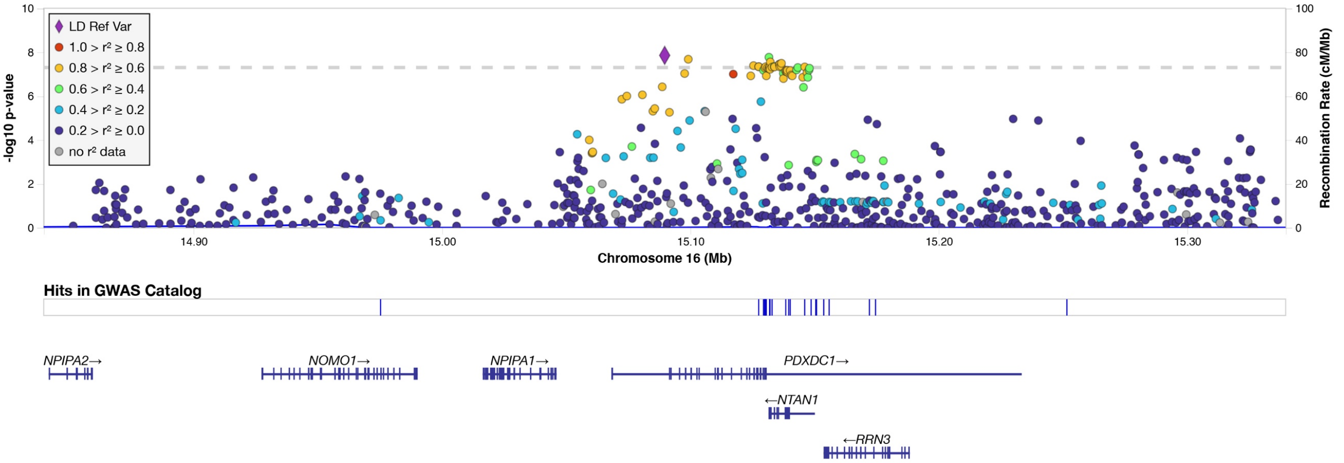

## rs6503451

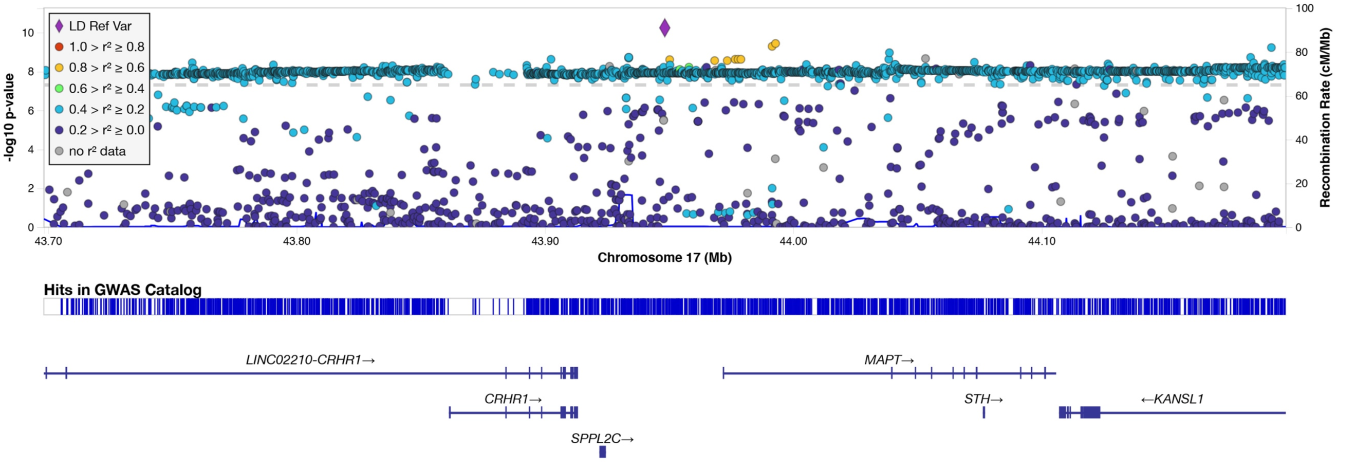

## rs199502

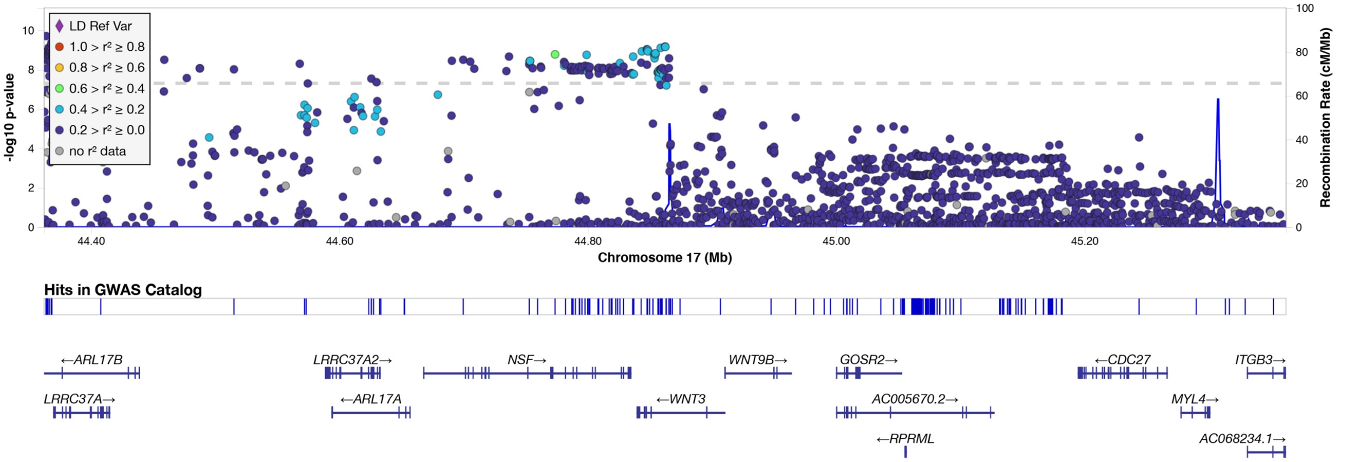

rs62621197

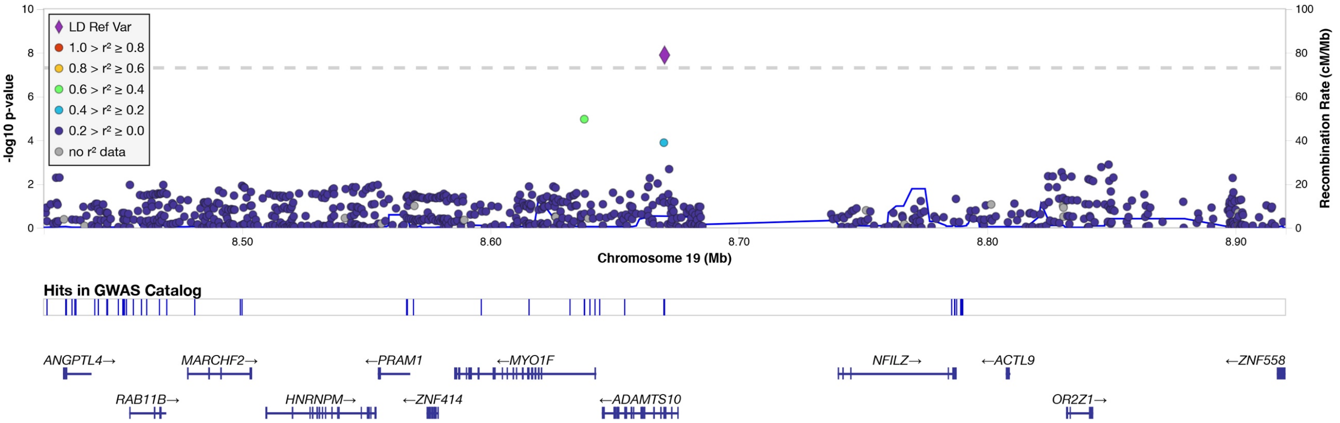

rs56252725 (EUR only)

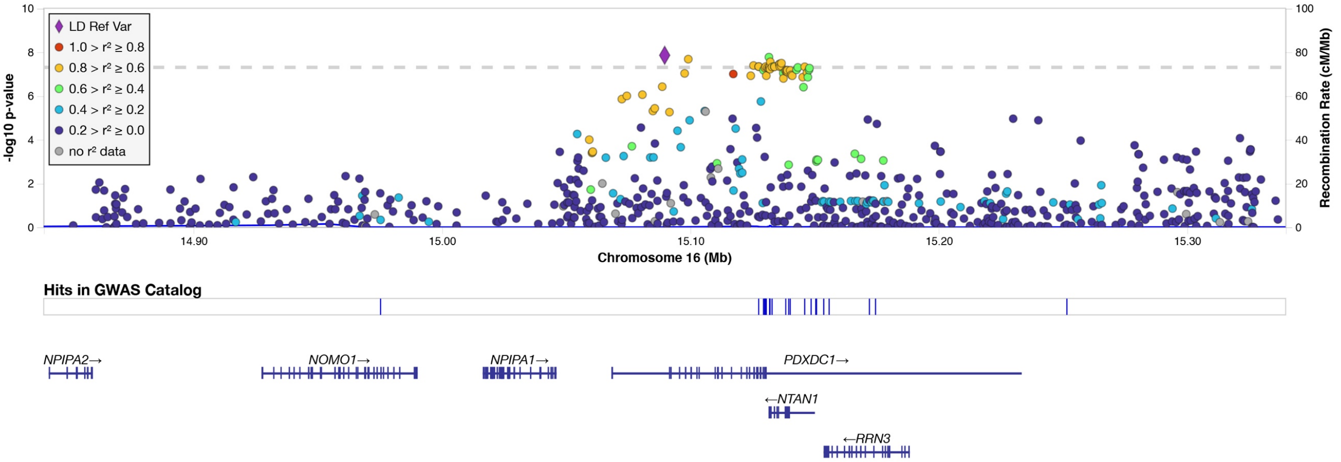

For each plot, the sentinel single-nucleotide polymorphism (SNP) is depicted by the purple diamond and labeled above each plot. Adjacent SNPs are depicted as circles, with the degree of linkage disequilibrium with the sentinel SNP shown by color (see legend). Genome-wide significance is shown by the hashed horizontal line. Genes in the region and their locations are shown at the bottom of each plot.

**Figure 4.** Manhattan plot of unindexed LV mass GWAS

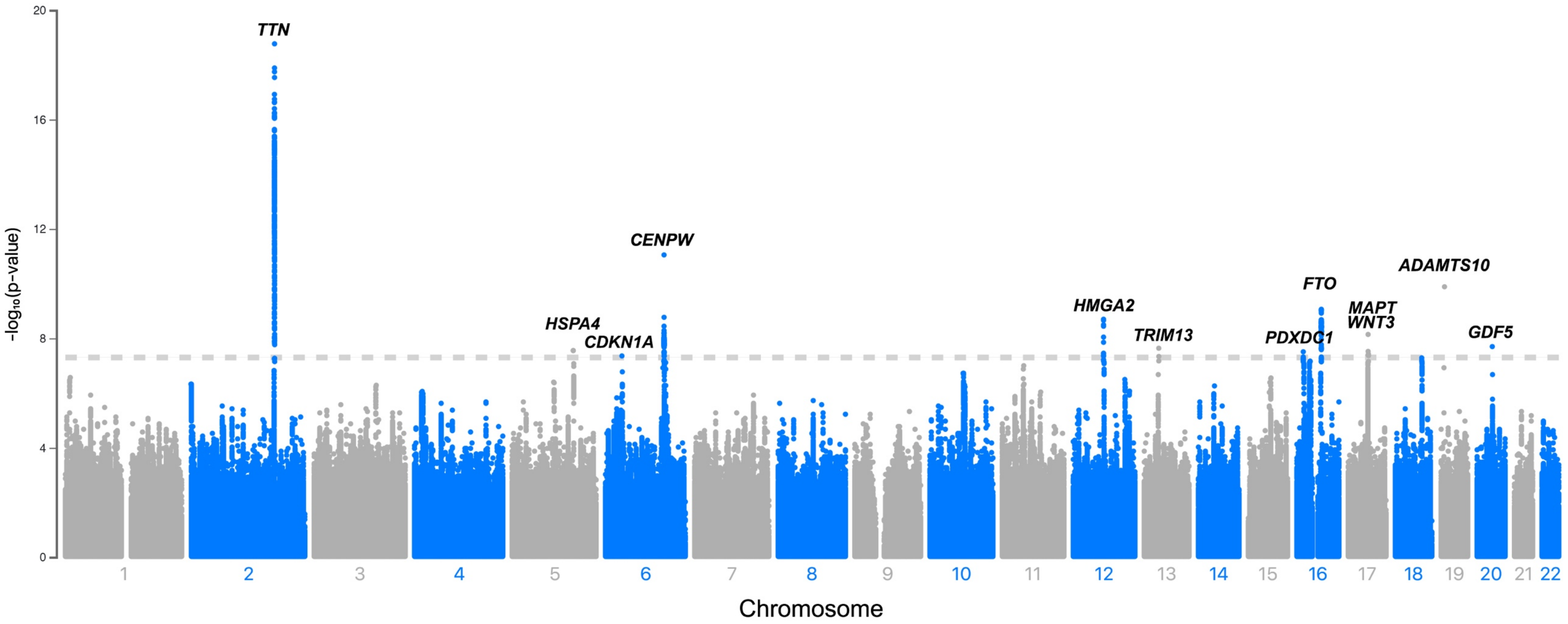

Depicted across increasing chromosome (x-axis) are the results of the European ancestry subset GWAS of left ventricular mass index. Variants meeting genome-wide significance ($5x10^{-08}$, depicted by hashed horizontal line), are labeled by the closest gene to the locus.

**Figure 5.** Quantile-quantile plot of LVMI GWAS for European-ancestry sample

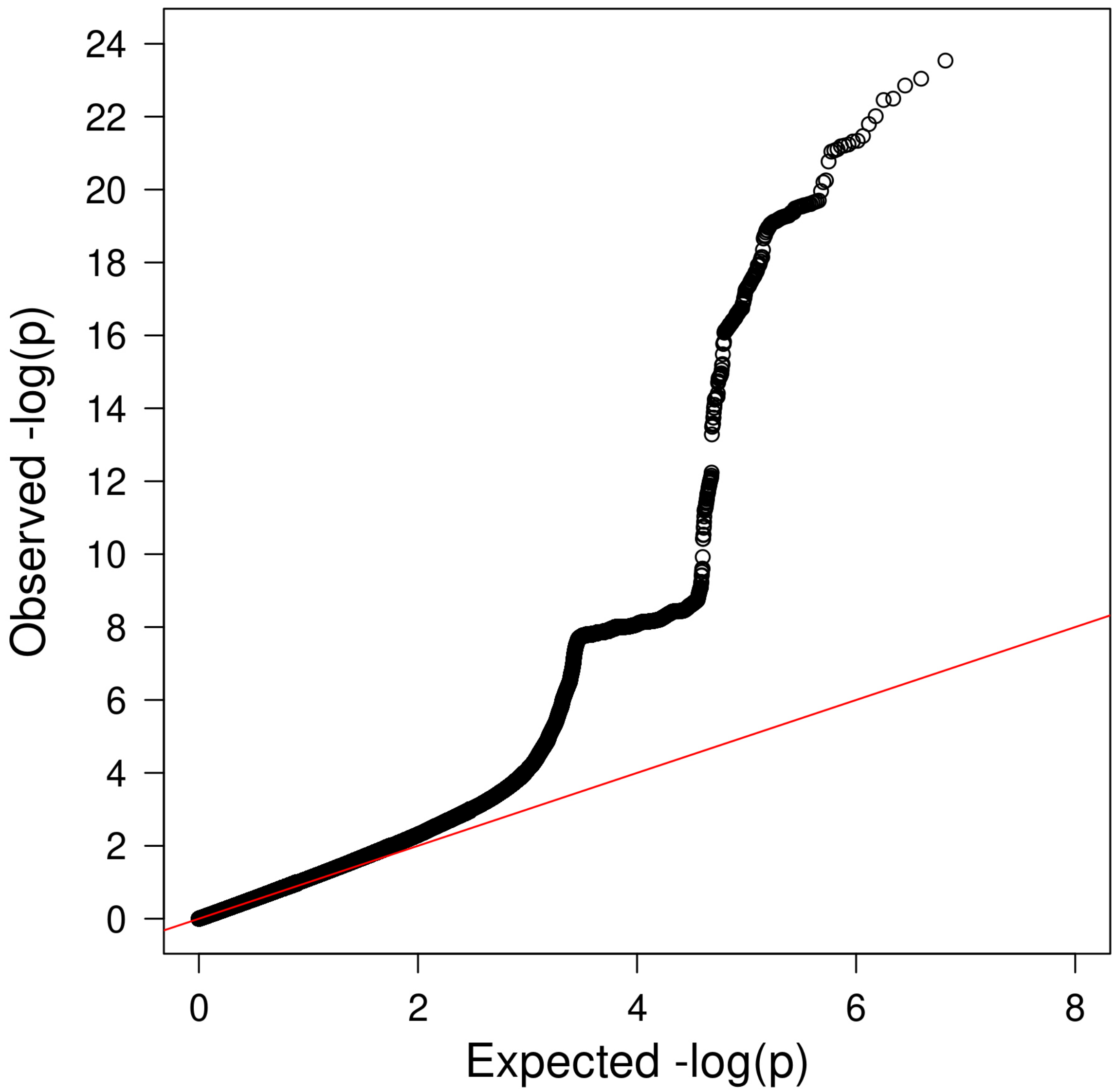

The lambda genetic control factor was 1.15. Linkage disequilibrium score regression analysis[3] revealed an intercept of 1.00, suggesting that the elevated lambda value was attributable to polygenicity rather than inflation.

**Figure 6.** Manhattan plot of European-ancestry GWAS

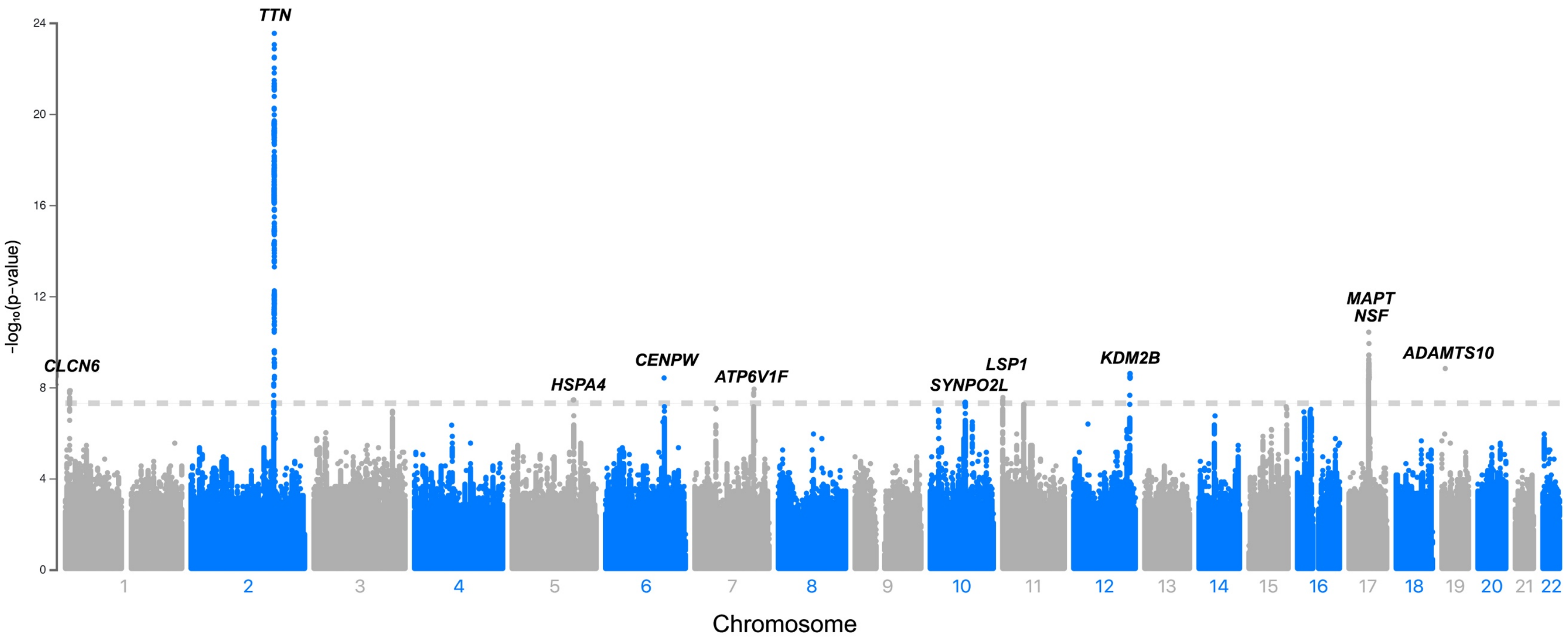

Depicted across increasing chromosome (x-axis) are the results of the European ancestry subset GWAS of left ventricular mass index. Variants meeting genome-wide significance ($5x10^{-08}$, depicted by hashed horizontal line), are labeled by the closest gene to the locus.
